# High-throughput Mutational Surveillance of the SARS-CoV-2 Spike Gene

**DOI:** 10.1101/2021.07.22.21259587

**Authors:** Ezgi Özkan, Marcus Martin Strobl, Maria Novatchkova, Ramesh Yelagandula, Tanino Guiseppe Albanese, Petr Triska, Lukas Endler, Thomas Penz, Timothej Patocka, Vera Felsenstein, Alexander Vogt, Ido Tamir, Tamara Seitz, Manuela Födinger, Ralf Herwig, Alexander Indra, Daniela Schmid, Christoph Bock, Andreas Bergthaler, Alexander Stark, Franz Allerberger, Ulrich Elling, Luisa Cochella

## Abstract

SARS-CoV-2 has evolved rapidly towards higher infectivity and partial immune escape over the course of the pandemic. This evolution is driven by the enormous virus population, that has infected close to 200 million people by now. Therefore, cost effective and scalable methods are needed to monitor viral evolution globally. Mutation-specific PCR approaches have become inadequate to distinguish the variety of circulating SARS-CoV-2 variants and are unable to detect novel ones. Conversely, whole genome sequencing protocols remain too labor- and cost-intensive to monitor SARS-CoV-2 at the required density. By adapting SARSeq we present a simple, fast, and scalable S-gene tiling pipeline for focused sequencing of the S-gene encoding for the spike protein. This method reports on all sequence positions with known importance for infectivity and immunity, yet scales to >20K samples per run. S-gene tiling is used for nationwide surveillance of SARS-CoV-2 at a density of 10% to 50% of all cases of infection in Austria. SARSeq S-tiling uncovered several infection clusters with variants of concern such as the biggest known cluster of Beta/B.1.351 outside Africa and successfully informed public health measures in a timely manner, allowing their successful implementation. Our close monitoring of mutations further highlighted evolutionary constraints and freedom of the spike protein ectodomain and sheds light on foreseeable evolutionary trajectories.

## Introduction

Within one year of pandemic, SARS-CoV-2 has diverged into a multitude of variants relative to the sequence first described to circulate in humans^1,2^. Selective pressure for changes that increase viral fitness has resulted in expansion of several SARS-CoV-2 variants of concern (VOCs) with complex sets of mutations^3^. Typically, amongst them are mutations that increase binding affinity to the receptor ACE2^4^ and/or mutations that at least partially evade the adaptive immune response to the initial strain^5,6^. The latter is of growing concern due to increasing immunity across the population, either from overcoming a prior infection or from vaccination^2^. Some of these variants have swept through the globe (B.1 and B.1.1.7=Alpha) or are dominating locally. Some variants or their derivatives are expected to globally spread in the future (B. 1.351=Beta, P.1=Gamma, B.1.617.2=Delta). Large second or third waves of infection have been observed as a consequence of increased infectivity and/or immune evasion. Monitoring the emergence and spread of such variants of concern is thus imperative for two reasons. First, to prevent or delay outbreaks of variants against which there is insufficient immunity in the population, by coupling variant detection to contact tracing and isolation. Second, identification of variants of concern is essential to inform the production of second-generation vaccines to curtail the spread of SARS-CoV-2.

Detection of many of the currently known mutations of concern in the SARS-CoV-2 genome can be done with specific quantitative RT-PCR approaches^7,8^. These can provide fast results, within a few hours from sample collection, and are thus an essential tool for rapid cluster isolation. However, such approaches do not allow for unbiased identification of novel mutations. Moreover, many of the current SARS-CoV-2 variants carry multiple mutations affecting viral biology, so differential diagnosis of all relevant combinations by RT-qPCR becomes impractical, error-prone, and often unfeasible. Genome-sequencing overcomes these challenges, albeit at higher costs, labor and time, in large part because most sequencing efforts have been focused on the complete genome of SARS-CoV-2. Various methods and toolkits are available to sequence the complete ~30 Kb genome of SARS-CoV-2 with next generation sequencing (NGS)^9–11^. These have been used to detect and monitor the rise of mutations in different countries^12–17^. For effective surveillance of SARS-Cov-2 variants, European Commission recommends sequencing of at least ~5% of total positive cases^18^. However, because of cost and ease of implementation, these methods have covered relatively small fractions of SARS-CoV-2 positive patient samples. Only small fractions of positive samples have been sequenced in some of the most severely affected countries like India (0.085%) and Brazil (0.095%)^19^. In the USA less <1% of samples were sequenced in 2020 when pandemic peaked. In the European Union countries, the fractions ranged between 70% and <0.06% e.g. in Hungary (0.0539%), Czech Republic (0.252%), and Poland (0.539%), with just a few outliers as Denmark (since Jan. 2021 sequencing >80% of cases) and UK (which went from ~10% in Feb. to >50% in March)^19,20^. Good mutational surveillance in most countries, including regions with high COVID-19 prevalence in Asia, Africa and South America is still lacking.

All mutations affecting infectivity of SARS-CoV-2 described to date, reside in the ectodomain of the spike protein. Whereas most of the protein shows high conservation among coronaviruses, the N-terminal domain (NTD) as well as the receptor binding domain (RBD) show a high degree of evolutionary flexibility (**Supplementary Fig. 1A**). The RBD is of evident importance as changes in this region can alter binding properties to the main receptor, ACE2, but possibly also to other host proteins^21,22^. In addition, relevant interactions with the human immune system also seem to occur within the NTD and RBD, as epitopes for neutralizing antibodies and for T-cell recognition reside within these regions^23^. Multiple mutations within these domains have been found to result in selective advantage and/or alter viral biology. In fact, all current SARS-CoV-2 VOCs are primarily defined by their complement of mutations in these regions of the spike protein. These include, among others, deletion of amino acids H69 and V70, S477N, N501Y, D614G, and P681H all of which have shown strong selective advantage alone or in combination^23^. In addition, most current vaccination efforts are based on the spike protein. Therefore, all mutations that can cause evasion of vaccine-induced immunity are anticipated to occur within the spike protein. Mutations L452R (found in B.1.617 among others) and E484K or E484Q as observed in strains Gamma (P.1), R.1, Beta (B.1.351), and B.1.617 as well as mutation of N439K (B.1.258), K417N (Beta) or K417T (P.1) are such examples^2,5,6,24^. For these reasons, variant surveillance sequencing efforts can be focused on the 10% of the SARS-CoV-2 genome encoding for the spike protein. The SARS-CoV-2 S gene, encoding the spike protein, spans 3819 nucleotides but the domains where most mutations of relevance have arisen comprises about two thirds of this. We thus sought to set up a high throughput sequencing pipeline focused on this region, spanning around 2100 nucleotides. Such a focused approach simplifies the procedure relative to whole genome sequencing methods and reduces reagent and sequencing costs as well as time of processing and analysis.

Here, we describe a simplified S-gene tiling approach using SARSeq as a high throughput, robust, and cost-effective workflow for sequencing the ectodomain of the SARS-CoV-2 spike protein for thousands of samples in parallel. We show that SARSeq S-tiling achieves full coverage of the region of interest in samples with Ct values as high as 33-35 and can reliably detect known and identify novel variants present in as little as 1% within one sample. We also benchmarked SARSeq S-tiling against whole genome sequencing (WGS) and further provide a simple bioinformatic pipeline to report sample-based results. We used SARSeq S-tiling for surveillance of 10-50% of all COVID19 cases in Austria during the period of January 2021-June 2021. SARSeq S-tiling uncovered clusters of various VOCs and multiple other mutations of interest and could thereby inform public health measures to contain these threats.

## Results

### Design of a robust, scalable and cost-effective mutation surveillance pipeline

Mutational surveillance to detect changes in the pathogens genome during a pandemic demands solutions that are both robust and simple to implement, but also scalable and cost-effective. Sequencing the genome of the pathogen of interest using NGS offers an optimal approach for standardized high-throughput analysis. However, several challenges must be met: **i)** the sensitivity of the analysis approach must be sufficient to deliver results across the range of viral titers observed in positive patients; **ii)** reliable scalability to thousands of patient samples as well as multiple amplicons per sample in parallel; **iii)** short turnaround time for sequencing results to empower case-based follow-up measures; **iv)** rapid setup of operations to enable quick responses; **v)** costs must be kept as low as possible as they might otherwise be prohibitory for large sample sizes. We sought to develop a simple and fast workflow for SARS-CoV-2 S gene sequencing that overcomes these challenges and enables the analysis of thousands of samples in parallel by tackling the rate-limiting steps of current WGS protocols: **i)** eliminating the need for labor- and time-intense NGS library preparation downstream of PCR amplification and **ii)** reducing sequencing time **iii)** increasing the scalability to thousands of samples in parallel. We started by designing a set of PCR amplicons that tile the desired portion of the S gene (**Fig. 1A, B**). The widely used ARTIC design for SARS-CoV-2 tiles the whole genome with ~400 bp amplicons to which sequencing compatible adapters are added by library preparation step and prepared library must be sequenced using a 500-cycle kit, typically in paired-end 250 modality (PE250) in case of Illumina platform^11,25^. We aimed to design a sequencing approach around a PE150 sequencing modality with amplicons of up to 281 bp, excluding adapters, to reduce costs, improve sequence quality, and increase sequencing speed for faster turnaround time and higher sequencing throughput per machine. This approach reduces sequencing time by about 12 hours and costs by EUR 1400/run in our setting. The PE150 modality further avoids competition with the standard WGS pipeline for potentially limiting reagents. Amplicons were partially overlapping in order to cover the whole region of interest, yet we minimized this overlap for most amplicons. A notable exception is the most variable region of the RBD^21^ from amino acid 470 to 502, where a 105 bp overlap was incorporated in order to cover this critical region, which harbors several known mutations of concern, with two independent amplicons (**Supplementary Fig. 1**). This design resulted in 12 amplicons that tile the N-terminal part of the SARS-CoV-2 S gene plus another amplicon covering a known T-cell epitope (**Fig. 1B**)^26–28^, thus allowing us to detect mutations affecting the protein surface with duplicate coverage of the most flexible region of the RBD.

**Figure 1.**
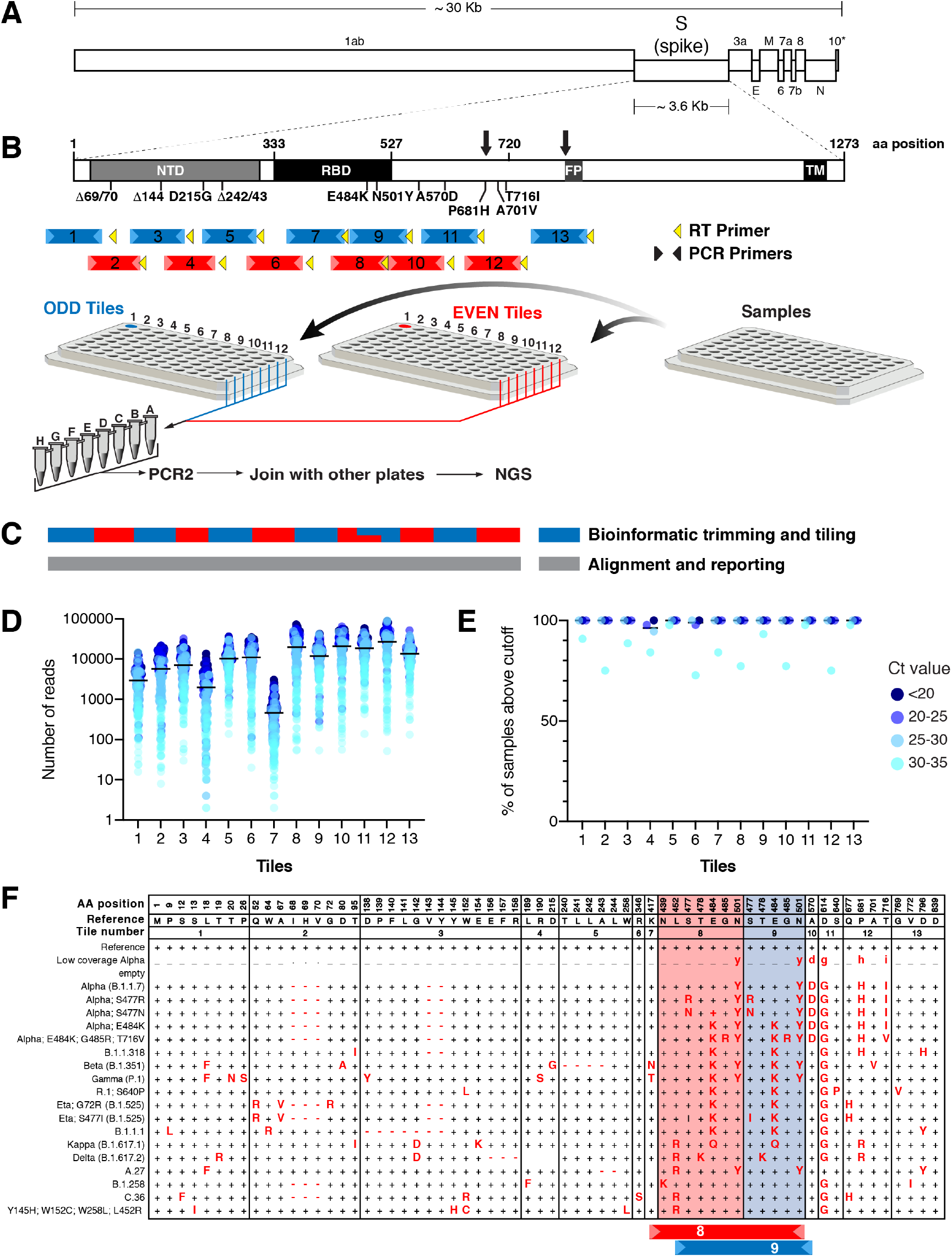
A SARSeq based pipeline for high throughput sequencing of the SARS-CoV-2 S gene. **A.** Schematic view of the SARS-CoV-2 genome. The S gene, coding for the spike protein, makes up about 12% of the viral genome. **B.** Linear scheme of the spike protein coding sequence. Known mutations of concern and with phenotypic consequences cluster in the first two thirds of the spike coding sequence including the N-terminal domain (NTD), the receptor binding domain (RBD) up to the protease cleavage sites (arrows) and fusion peptide (FP). The subsequent stalk region and transmembrane domain (TM) is mostly devoid of such mutations. Tiles to cover the spike coding sequence from amino acids 1-722 and 767-839 are colored blue (Odd tiles) and red (Even tiles), primers used for amplification are highlighted in lighter color. Tile 13 is non-continuous with tiles 1-12 and covers a known T-cell epitope26-28. Tile specific RT primers are shown as yellow triangle. Odd and Even reactions are performed separately for RT and PCR. Plates are subsequently pooled to single columns, where Odd and Even tiles are united, and a single PCR2 reaction is performed for pools of 12 patient samples. All reactions are then combined, gel purified, and sequenced on an Illumina NovaSeq platform. **C.** Overlapping termini of tiles are resected bioinformatically. The intentional large overlap between Tiles 8 and 9 over the RBD is reported twice independently. Each fragment is aligned to the consensus sequence and the all fragments are stitched together for the final output. **D.** Read counts obtained for each Tile, for 179 samples ranging from Ct=14.9 to Ct=34.8. Ct ranges are indicated with color code. **E.** Fraction of samples with successful amplification is shown for each Tile. Amplification success is determined based on the implemented cutoff (1% of reads relative to median of the tile and a minimum of 10 reads). Samples are binned by Ct ranges, as indicated by the color code. **F**. The complete sequence is reported as table. Reference positions are denoted as “+” or “_” if below confidence cutoff (see text and **Fig. 2**). Amino acid changes are in uppercase for positions with >50% of reads being mutant, but 20%-50% of mutant reads or mutations below cutoff are shown in lowercase. Deletions are denoted by “−” or “.” if below cutoff. Blank cells indicate no reads. Positions 370-502 are reported in duplicate based on the independent amplicons 8 and 9.

To implement a simple, cheap and high-throughput workflow for amplification and sequencing we adapted SARSeq, a method we recently developed for detection of SARS-CoV-2 and other viruses^29^. SARSeq is a robust pipeline to reverse transcribe and amplify multiple nucleic acid segments in parallel, and across thousands of individual samples, while maintaining high sensitivity and specificity. SARSeq uses a two-dimensional, unique dual indexing strategy, in which every sample is barcoded directly in two subsequent PCR reactions with 4 indices in total. By also incorporating all required adapter sequences (i5, i7, p5, p7) in these steps, the PCR products generated by SARSeq can be directly sequenced by Illumina platforms, without the need for further library preparation steps^29^. This workflow reduces the entire process to 6 pipetting steps where reagents are added or pooled and does not require any intermediate purification step (**Supplementary Dataset 1 and Methods**). In brief, these steps are **i)** addition of sample to the RT mix, **ii)** addition of PCR1 mix to the RT reaction where sample-specific indices are added, **iii)** pooling, **iv)** addition of ExoProStar to remove unused primers, **v)** addition of PCR2 mix where a second set of indices for combinatorial barcoding are added, as well as adaptors for Illumina sequencing, and **vi)** final pooling.

To achieve sensitive, robust and uniform amplification across the S gene, multiple experimental parameters had to be adapted relative to the original setup^29^. First, because overlapping amplicons cannot be produced within the same reaction, we split the “Odd” and “Even” amplicons or tiles in two independent RT- and PCR reactions using the respective primers for each set (Fig. 1B), as is also done for whole genome sequencing using the ARTIC primer design^25,30^. For each sample, both the Odd and Even PCR products receive the same index pairs. Second, we found that using a specific RT primer with a melting temperature of ~58°C for each amplicon, as opposed to conventional random priming using hexamers, resulted in markedly increased read counts for all amplicons (**Supplementary Fig. 2A, B**). In particular for Tile 7, that reproducibly gave the lowest read counts, the addition of a second tile-specific RT primer made a significant difference (**Supplementary Fig. 2C**). We assume that the longer gene-specific primers can still bind the RNA template at the RT temperature of 55°C, as opposed to hexamers, and thus support reverse transcription in regions with higher degree of secondary structure. Third, RT- and PCR protocols were adapted to suit the longer amplicons and suppress generation of short, unspecific amplicons (**Supplementary Dataset 1 and Methods)**. Fourth, RT- and PCR primer concentrations were optimized to balance coverage across tiles. Last, the general pooling scheme and setup of PCR2 was adapted to the use of many parallel amplicons (**Methods**). Together these adaptations yielded a robust PCR pipeline that successfully amplified all tiles in a multiplexed PCR reaction.

### Measures to ensure quality and specificity

When processing thousands of positive samples, cross-contamination is a major concern. We implemented the following measures to prevent sample cross-contamination: **i)** strict local separation of pre- and post-PCR steps, **ii)** addition of dUTP to both PCR reactions as well as UDG prior to starting PCR1 in order to degrade previously-amplified, contaminating DNA molecules, **iii)** use of distinct PCR1 indices in neighboring pools, **iv)** use of alternating PCR2 indices each week, and **v)** strict monitoring of undesired amplification products. To assess the effectiveness of these measures, we analyzed the number of reads per tile for a set of 627 wells, across multiple plates, that were expected to be empty based on information from the various diagnostics labs across Austria that provided the samples. 171/627 samples had zero reads across all amplicons, 277 samples produced a maximum of one or two reads for at least one amplicon, 143 produced between 2-9, and only 24 samples contained an amplicon with 10 reads. Low read counts are most likely due to index hopping^31,32^ that can occur at such low levels even when using unique dual indexing, when working with so many positive samples. To filter against spurious reads generated by index hopping, we implemented a cutoff for each tile, whereby we demanded at least 1% of the median read number of the respective tile in the run (or a minimum of 10 reads). This cutoff correctly eliminated spurious signal in 98.6% of presumed empty wells. In 9 wells (1.4%) however, we observed that samples appeared positive, showing amplification of >5, or often all tiles well above the defined cutoffs, indicating contaminating presence of SARS-CoV-2 RNA in those wells and not a false positive from our pipeline **(Supplementary Fig. 3**), corroborated by a mean read count of 13,198 per tile across those samples. Wells that we left intentionally empty in experimental plates prepared in our research lab did not show this problem, but rather we find this is more likely to happen in sample plates from diagnostic labs that handle very large numbers of positive samples. The defined per-tile cutoff was also used to derive an overall sample filter that reflected the overall quality of a sample: samples with fewer than 6/13 tiles above cutoff, were considered of insufficient quality and were not subsequently analyzed for variants.

### Pipeline sensitivity and scalability

Positive samples for SARS-CoV-2 typically span a large range of viral titers, with differences of up to six orders of magnitude^33^. If multiple samples are to be sequenced in a highly parallelized setting, each sample should produce read numbers where the range of 10^6^ is condensed to 1-2 orders of magnitude at most to avoid that high titer samples are over-represented and lower titer ones missed. To dampen this difference, we allowed PCR1 (on individual samples) to reach end-point. This blunted original differences in titer sufficiently to maximize the use of available sequencing depth.

We tested this optimized setup on a set of 179 samples ranging from Ct=14 to Ct=35. Amplification across all tiles was robust, yet average read counts varied between 456 (tile 7) and 26,880 (tile 12) reads (**Fig. 1D**). Using our established cutoffs, we observed that for each tile, 93%-99% of samples generated usable/reliable sequencing data (**Fig. 1E**). Sequencing results are reported as a table with reference positions denoted as “+” and mutant positions as the respective nucleotide or amino acid symbol. Viral variants carrying multiple mutations were summarized as the respective lineage (**Fig. 1F**, for further details on results output see Methods).

The ability to obtain sequencing data even from low titer or bad quality samples is an important feature of such a surveillance pipeline. To assess sensitivity directly, we correlated SARSeq S-tiling performance to Ct values measured with a commercial kit on the N1 amplicon^34^ for a set of 192 clinical samples (**Fig. 2A**). We found that all amplicons could be detected for 93% and 89% of samples at below Ct=33 and 35 respectively, which corresponds to <10 molecules per reaction^29^. The redundantly covered variable region of the RBD was detected in 99% of samples at these cutoffs (**Fig. 2B**). The RT-PCR protocol employing SARSeq S-tiling is thus efficient to amplify all tiles at similar sensitivity down to a few molecules per sample.

**Figure 2.**
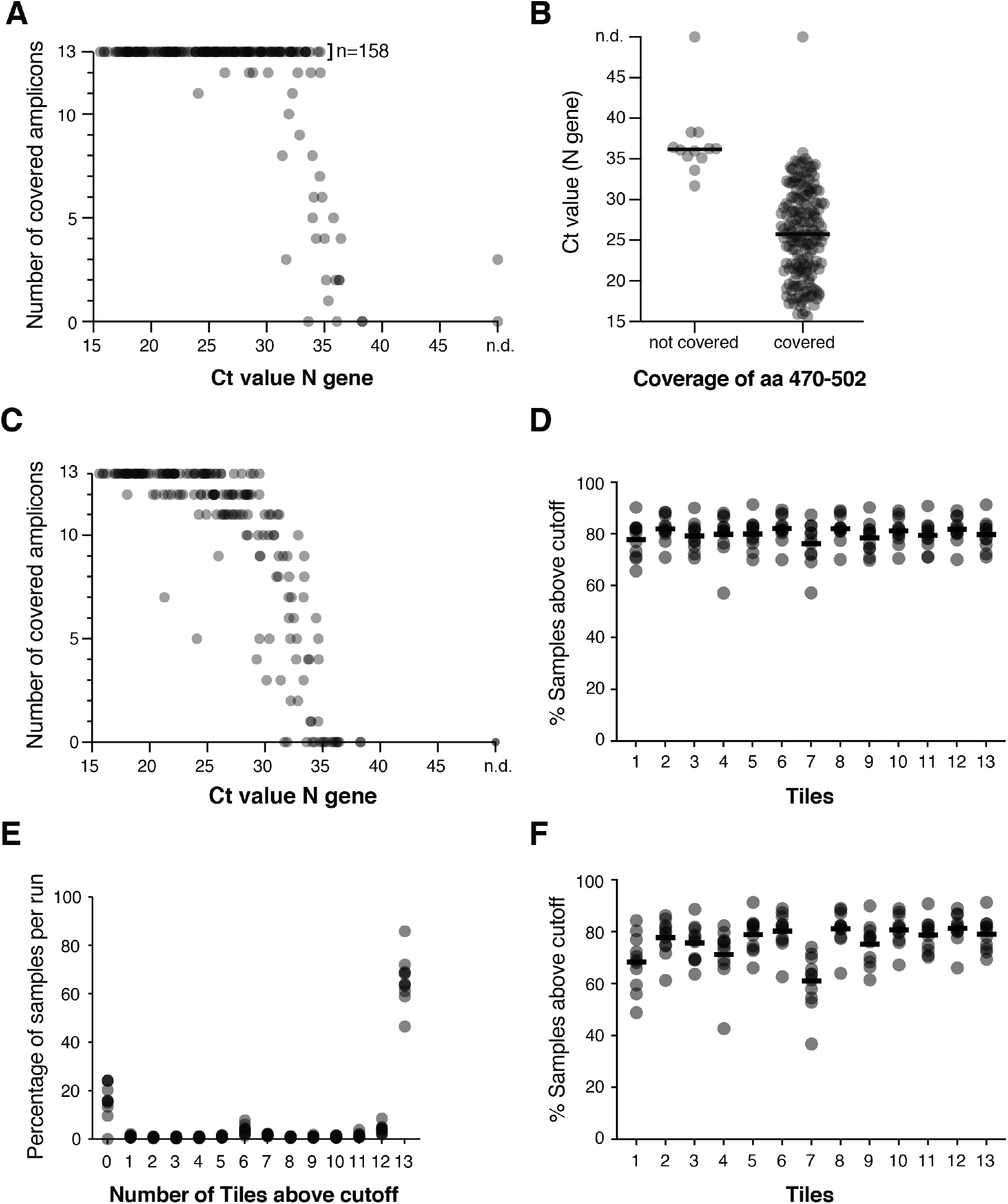
Amplification performance and sensitivity. **A**. Number of amplicons detected relative to Ct value. Most amplicons are recovered up to Ct~34-35. **B**. Coverage of the most flexible region of the RBD including most ACE2 interacting positions relative to Ct value. The region is considered covered if Tile 8 or Tile 9 (or both) are detected. **C**. Bioinformatic scaling of recovered amplicons to a hypothetical run of 23,000 samples in a single sequencing lane while maintaining the cutoff rules. Recovery remains near complete at Ct <30. **D**. Percentage of samples with successful detection of each of the 13 tiles above a confidence cutoff of >1% of reads relative to the median reads for that tile obtained in the run (or a minimum of 10 read). Data for 11 independent runs (each with 2,300 samples) is plotted. See Supplementary Table 1 for source data. **E**. Distribution of the number of detected amplicons per sample. Data for the same 11 runs as in D is shown. Samples typically generate all or no amplicons due to the even sensitivity across the covered region. **F**. Bioinformatic scaling of real-life sample data to hypothetical 23,000 samples analyzed on one flow cell while maintaining the cutoff rules. Even under these conditions, most tiles are covered in >70% of samples.

The sensitivity of any sequencing pipeline is inversely proportional to the number of samples that are pooled in one sequencing run. We therefore wanted to estimate how many more samples we could pool in one sequencing lane in the current setup (one NovaSeq lane, about one billion reads) and still maintain a similar level of sensitivity. To this end we down-sampled the reads from a typical run by a factor of ten and plotted the number of detected amplicons while maintaining the established cutoffs. Of the samples with Ct values <33 and <35 we could still detect at least 12/13 amplicons for 91% and 86% respectively (**Fig 2C)**. Tile 7, which produces the lowest number of reads, was still detected in 77% and 70%, and importantly, the variable region of the RBD was still covered for 99% of samples. Thus, the robust performance of the pipeline should allow for multiplexed sequencing of up to 23,000 patient samples with Ct<33 together in a single NovaSeq lane with a target output of 1 billion reads. This would bring sequencing costs to a few cents per sample and highlights the cost-effective and scalable nature of SARSeq.

### Setup of a surveillance pipeline for Austria

To set up nationwide mutational surveillance of SARS-CoV-2 in Austria we established a pipline that would process twenty-four 96-well plates weekly with a single NovaSeq PE150 run. To guarantee delivery of robust sequencing results we developed a standard operating procedure including robotic assisted pipetting, an optimized NGS read mode, and a hand-over protocol to initiate NGS analysis automatically upon completion of sequencing (**Supplementary Dataset 2**). We processed 24 x 96-well plates of positive samples, namely 2,304 wells, per week with a team of two full-time and two part-time members and delivered SARS-CoV-2 spike gene sequences from RNA preparations. This represented 10-50% (depending on SARS-CoV-2 incidence at the time) of all positive samples in Austria in the period of January-May 2021. Each processing cycle is initiated on Monday and sequencing data are delivered on Friday of the same week. Thereby, SARSeq could cover the required depth of mutational surveillance by sequencing suggested by the European Center for Disease Control for Austria.

To aid rapid bioinformatic analysis, we implemented an expert system for semi-automated and assisted manual annotation. In order to detect and quantify known and novel sequence variants of the SARS-CoV-2 S gene, the bioinformatic analysis was performed in several steps: i) the paired-end reads were assigned to sample IDs via the two-dimensional dual indexing as previously described^29^; ii) the reads of each sample were assigned to the intended tile via the corresponding primer sequences at the ends of each amplicon (invalid amplicons were discarded); iii) the amplicon sequences without the primers were aligned to the respective segments of the SARS-CoV-2 S-gene reference using a strategy that allowed for mismatches during alignment, and such variants were subsequently detected and quantified at the DNA and protein level (**Methods and github link**). Results are output as tables for manual inspection and annotation with each row showing one sample and each column showing one nucleotide or amino acid. Positions that change relative to the reference are highlighted for convenience. We also automatically generate a simplified version of the output table that only shows amino acids that differ from the reference and groups the samples by similarity of mutational signature. To facilitate variant annotation, samples are clustered based on mutational signatures (**Fig. 1D, Supplementary Dataset 3**). In addition, for each sample we provide the numbers and percentages of mapped reads per tile as well as other metrics to assess sample quality. The complete bioinformatic analysis takes ~5 hours to process 1 billion reads and 2,304 samples, producing an ideal output for manual sample annotation and case follow up.

For the Austrian surveillance pipeline, SARS-CoV-2 positive samples were delivered to us upon RNA purification as separate 96-well plates by labs throughout Austria. Despite deemed positive, the samples spanned the natural range of viral RNA titers and even included empty wells and (partially) degraded samples or with little remaining volume. We evaluated the performance of SARSeq S-tiling on ~25,000 samples, processed in eleven independent runs. We observed that all tiles were relatively uniformly covered in ~80% of samples, using the cutoffs described above (Fig. 2D). Failed tiles were not uniformly distributed across samples, but rather, samples tended to either produce reads for all tiles or for none (**Fig. 2E**), likely indicating low level, low quality or absence of any template RNA in ~20% of the samples. This is consistent with the amplicon-coverage for samples with low viral titers (Fig. 2A, C) and suggests that in a real-world setting, up to 20% of PCR-positive samples collected from various diagnostic labs are not compatible with sequencing.

We scaled these data to mimic the pooling of 23,000 samples per run (**Fig. 2F**). This would have reduced the performance of amplicons 1, 4, and 7 but still maintained a robust pipeline delivering sequence information for relevant regions of the S gene. SARSeq based S-gene tiling is thus sensitive to a Ct value of 35 and reaches good performance on real-life clinical samples across multiple sample providers.

### Pipeline Reproducibility

The next aspect we explored is the accuracy of the sequence information derived by the pipeline at the cutoffs we had set. To test reproducibility of our sequencing calls, we took advantage of the redundancy built in by our primer design. Specifically, amino acids 470-502 are covered by amplicons 8 and 9, which are processed in two independent reactions, the Even and Odd batch respectively. As multiple samples carry amino acid substitutions in positions S477, E484, and N501, we compared the calls for these positions from tile 8 and tile 9 for >10,000 samples. We found a near perfect agreement for samples where data was available from both amplicons (**Fig. 3A**). Both the reference sequence as well as mutations are called with extremely high reproducibility. Only two cases where observed, where a high confidence mutation is not confirmed in the other amplicon and might represent a PCR- or sequencing error. The error rate is thus 2 out of 3 x 10,000 or <0.01% per amino acid. Mutation calls with low confidence (from read numbers between 10 and the median-based read cutoff) and/ or sub-stochiometric representation within the sample (i.e. changes reported in lowercase) show a slightly lower coverage reproducibility but no discrepancy between results (**Supplementary Fig. 4A**). The error rate we calculated estimates that ~1/20 samples will have one error across the covered ~700 amino acids. We therefore placed an additional cutoff, and only report amino acid changes when they occur in three or more samples within each batch of 2,304 samples we process together. The probability that identical changes in any amino acid occur with this frequency is infinitesimal, thus we chose this stringent cutoff to report variant positions. SARSeq S-tiling therefore generates high-confidence sequence analysis of most of the SARS-CoV-2 S gene at low cost and high throughput.

**Figure 3.**
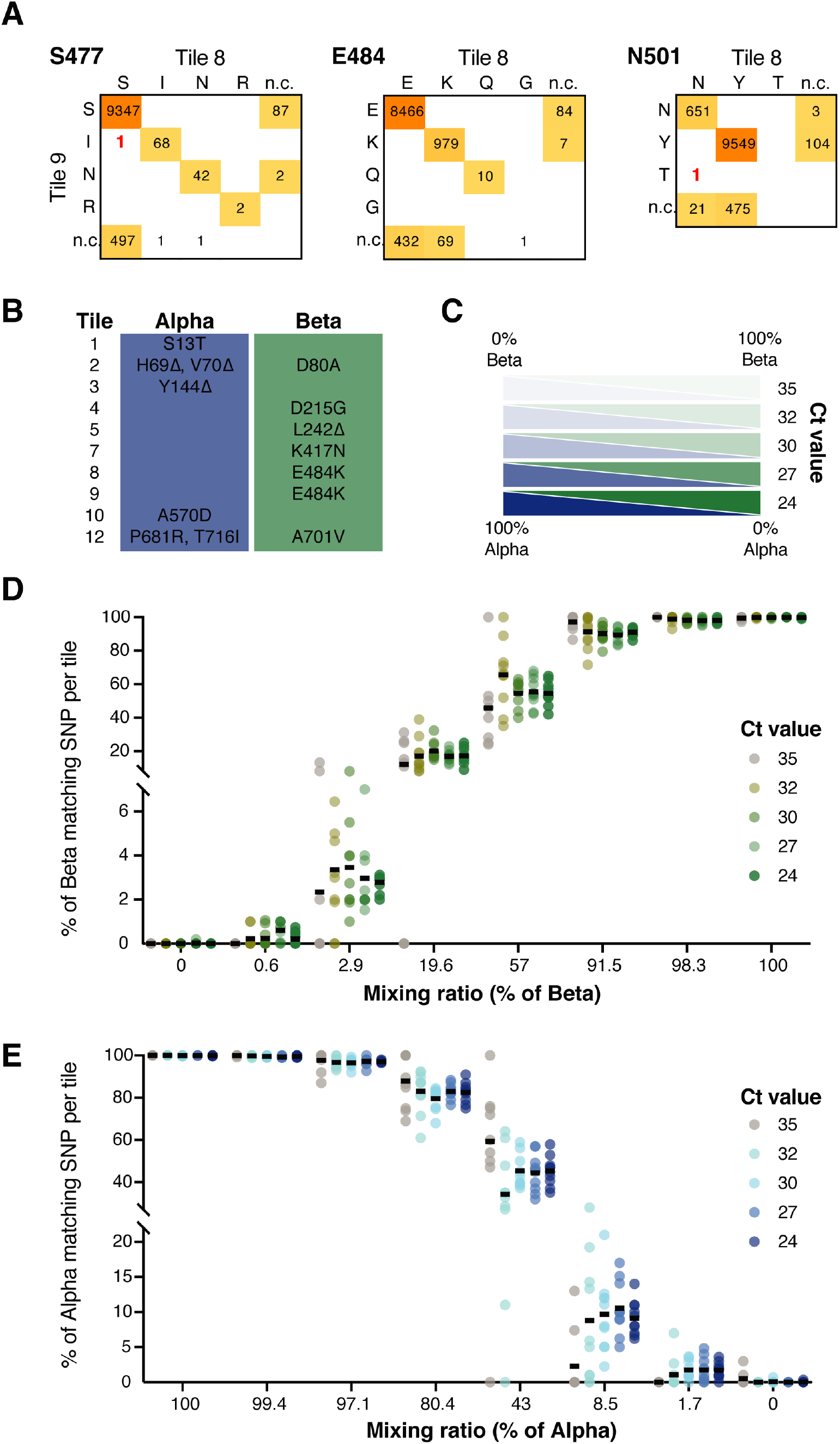
Reproducible, sensitive and quantitative detection of different SARS-CoV-2 variants. **A**. Sequence concordance between amplicons 8 and 9 across >10,000 samples, see also Supplementary Fig. 4A. Columns: results based on amplicons 8, rows: results based on amplicon 9. Two cases of discrepancy within the >30,000 datapoints (samples x positions) are highlighted in red. **B**. Discriminating positions between Alpha with S13T and Beta variants in 10/13 tiles. **C**. Graphical illustration of the experimental outline of mixing two distinct strains at different ratios and a subsequent dilution series. **D**. Fraction of reads indicative of the Beta variant in dependence of Ct values and mixing ratios. Individual datapoints refer to each of the 10 tiles used for the analysis. **E**. Same as in D but for the Alpha variant. As expected, differences across tiles increase with higher Ct values and sub-stochiometric variants are sometimes not detected at high Ct value. See Supplementary Table 2 for source data.

We finally challenged the sensitivity and robustness of the pipeline to call mutations present at low frequency within a single sample. To do this, we selected two variants that differ in 10/13 tiles, namely Beta and Alpha harboring an additional S13T mutation (**Fig. 3B**). Purified RNA from patient samples was mixed at different ratios and a dilution series was generated from these mixes to a Ct range of 24-35 as confirmed by RT-qPCR (**Fig. 3C**). Samples were then subjected to SARSeq S-tiling alongside a batch of 2,300 other samples (**Fig. 3D, E**). For every condition, we extracted the percentage of each of the two variants in every individual tile. SARSeq S-tiling quantitatively retrieved the fraction of each variant in the respective mixes. Concordance across tiles was high even for the more diluted samples, with standard deviations between amplicons in equimolar mixes ranging from 7% at Ct=24 to 28% at Ct=35 (**Supplementary Fig. 4B**). SARSeq S-tiling was even able to robustly quantify a spike-in of 1% of a virus strain in the background of a different strain. Whereas this was beyond the goal of our study and our surveillance responsibilities, such low-frequency mutations were indeed frequently observed and can be used to gain further insight into lineage relationships between sequenced samples^15^. In summary, SARSeq S-tiling delivers qualitatively and quantitatively reproducible sequence across the ectodomain-coding region of the S-gene, that contains most mutations currently considered to be biologically impactful.

### Benchmarking to WGS

To further benchmark the output of SARSeq S-tiling to an established WGS protocol^25^ using the ARTIC primer set, we processed 159 samples in parallel by both pipelines. We extracted the consensus sequence for each sample as obtained by the two pipelines, and performed pair-wise alignments (**Fig. 4A**). Of the total of >125,000 positions (795 aa x 159 samples) covered by both methods we found concordant results for ~92%, whereas for 7.9% of positions one or both methods lacked sequence coverage. Only 30 positions (0.02%) showed discrepancies between the two reported consensus sequences (**Fig. 4B**). These mismatches were not randomly distributed but rather found exclusively in positions where mutations are frequent: out of the 795 covered amino acids, only 178 are frequently substituted/deleted (cutoff at >4 mutations across all samples), but the 21 discrepant mutations where SARSeq called a mutation all correspond to such frequently mutated positions. This strong enrichment (4.4-fold; hypergeometric p-value=9.7×10^−15^, **Fig. 4C**) indicates that discrepancies are not due to random sequencing or analysis errors, but rather have three possible explanations: i) human error/contamination in handling samples and ii) RNA viruses exist as heterogeneous “quasispecies” with many mutations being partially penetrant in an individual sample; this, combined with amplification ratios could result in different calls from two independent analyses. iii) coinfection of an individual with genetically distinct viruses as previously postulated^35^. In support of that, for 6 mutations where discrepant calls are made we indeed see the mutation in a fraction of reads (11-78%), making it possible that independent PCR reactions bias towards one or the other variant in the mix and the most abundant one is used to extract the consensus sequence. Nevertheless, because of this extremely low frequency of discrepancies and the fact that they arise from reasons beyond the pipeline, we conclude that SARSeq S-tiling provides results that are in good concordance with WGS over the entire covered region.

**Figure 4.**
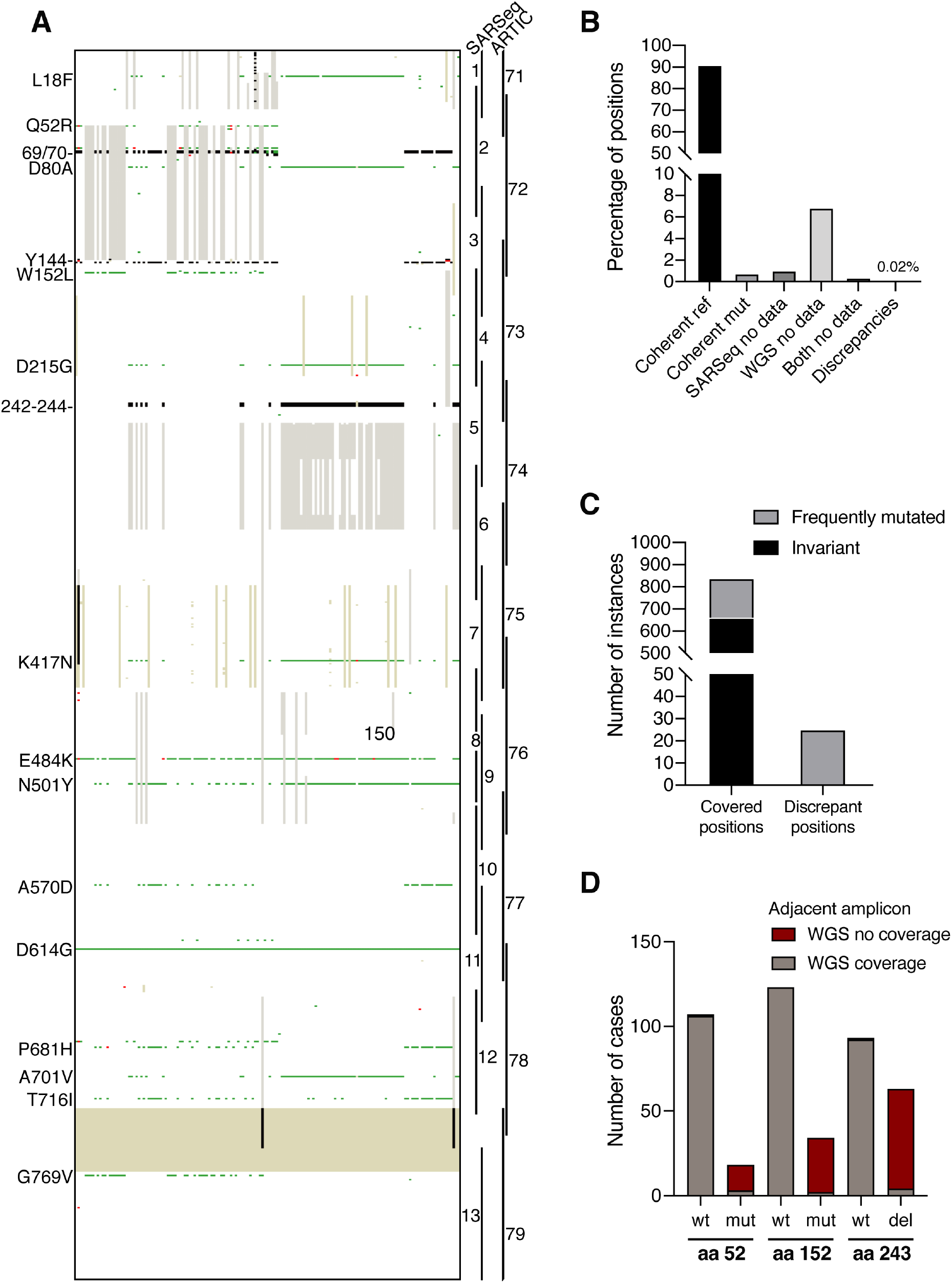
Concordance with whole genome sequencing. **A**. Heat map illustrating agreements and discrepancies between whole genome sequencing using the ARTIC primer set and SARSeq S-tiling. Each column is one sample, each row is an amino acid position. White: concordant reference call; green: concordant mutant call; red: discrepant call, gray: sequence missing in WGS; beige: sequence missing in SARSeq. The sequence coverage gap between SARSeq tiles 12 and 13 appears as solid beige block. Black: sequence missing in both datasets including genetic deletions e.g. in positions 242-244. **B**. Quantification of each classification as in A. A total of 30 discrepancies was found accounting for <0.02% of compared positions. **C**. Number of amino acid positions showing change in the entire dataset of 30,000 samples (Compare to Fig. 6C) or among the 25 discrepant positions where SARSeq calls a mutation (for the other 5 discrepancies, WGS calls a mutation but SARSeq finds the reference nt). Whereas only 179/795 of the analyzed positions show change, all 25 discrepancies occur in variable positions. Mutations are thus non-randomly distributed (see also Fig. 4A) and therefore not generated by the sequencing pipeline but a feature of the samples analyzed. **D**. Number of analyzed samples with or without sequence coverage of the amplicon neighboring to mutations in positions 52, 152, and 243. Amplification of neighboring amplicons fails when these mutations are present as they affect the primer binding site (see Supplementary Fig. 5).

The pair-wise alignments of sequences produced by both pipelines highlighted a patterned lack of sequence coverage, in which failure of an amplicon coincided with a mutation in the vicinity (**Fig. 4A**). Specifically, we noticed a failure of ARTIC tile 71 in samples carrying Q52R mutations, of tile 72 in case of W152L, and of tile 74 in case of deletion of 242-244 (**Fig. 4D**). Such correlations can be explained by the overlap of these mutations and the respective primer binding sites. We therefore asked how frequently would mutations affect the efficiency of amplification of different tiles. We extracted the most frequent mutations observed during our surveillance efforts from ~45K cases in Austria (43 mutations found in >100 samples) and asked how many overlap with either ARTIC and SARSeq S-tiling primer binding sites (**Supplementary Fig. 5A**). Indeed, the observed ARTIC amplicon failures we had observed are well explained by mismatches close to the primer’s 3’ end. In addition, one relatively frequently observed mutation (9.6% of the Austrian surveillance cohort), N439K, likely affects efficiency of the ARTIC amplicon 75, but might also affect SARSeq amplicon 7, which uses a 3’ extended primer. We therefore correlated read depth of SARSeq amplicon 7 to the N439K mutation. Satisfyingly, we did not observe reduced efficiency dependent on the mutation. In contrast, M177I, which is rare in our dataset (0.52%), generates a mismatch in the 3’ end of the SARSeq S-tiling reverse primer 3 and expectedly inhibits amplification. Adaptations of primer sequences might therefore be required as the mutation landscape of SARS-CoV-2 continues to evolve. In fact, an alternative ARTIC primer 72R circumventing the inhibition by W152L was already proposed^10^. Amplicon failure due to acquired mutations is thus observed in several instances and primer performance must be closely monitored.

### Results from nationwide mutation surveillance in Austria

We used SARSeq S-tiling in a high throughput effort to monitor the mutational landscape of SARS-CoV-2 in Austria. Positive SARS-CoV-2 samples were collected from all regions of the country beginning in January 2021 and, for this publication, until May 2021. This allowed us to systematically monitor prevalence and dynamics of mutations in space and time. Due to the limited sequence coverage, tiling of the S-gene is insufficient to enable robust automated lineage identification using the Pangolin software package^36^. We thus employed a custom solution for calling strains based on clustering and manual inspection. For all samples harboring at least one mutation seen in >3 samples we reported the full mutational fingerprint within the analyzed region. Complex sets of mutations such as those found in Alpha, Beta, Gamma, Delta, Kappa (B.1.617.1), Eta (B.1.525), B.1.258, R.1, and others, yielded sufficient certainty to be assigned a lineage annotation.

The Austrian state of Tyrol was of particular interest as by early January, it harbored multiple VOCs, providing a view of the relative fitness of several variants side-by-side, over several months. Within 8426 analyzed samples from January to May 2021 we detected 6048 cases of Alpha, 535 cases of Beta, 527 cases of Alpha with an additional mutation at position of E484K, and various other frequent mutations, most notably S477N and N439K (B.1.258 and others) (**Supplementary Table 3**) and were able to track the frequency of these variants across time (**Fig. 5A**). At the beginning of January, half of the cases in Tyrol were attributed to the reference SARS-CoV-2 strain, yet this disappeared by calendar week 9 (early March). The first VOC to expand in Tyrol during this period was Beta, which originated in South Africa. Initiating from Zillertal, a ski resort valley, Beta expanded from 9% in Week 2 to 30% in week 6 likely due to its higher infectivity relative to the reference strain. With a total of 535 sequence verified samples during this time period it represents the biggest reported outbreak of Beta outside South Africa. Similar to the reference strain and other circulating variants at that time, the incidence of Beta declined in the weeks after, as the incidence of Alpha steeply increased in Tyrol, as in the rest of the country (not shown). Importantly, an Alpha sub-strain harboring an additional E484K mutation, initially still expanded at a similar rate as Alpha in calendar weeks 9-11, and reached a relatively constant fraction of 7-10% of samples since. These results confirm the well-described increased relative fitness of the Alpha strain, supporting the notion that it outcompetes other VOCs such as Beta. Mutations in E484K have been associated with decreased effectiveness of neutralizing antibodies. The relative fitness of Alpha to its variant with E484K and relative to Gamma (with N501Y and E484K mutations) and Delta (lacking N501Y but displaying increased infectivity^37^) remains to be observed closely, in particular with an increasing fraction of the population being vaccinated.

**Figure 5.**
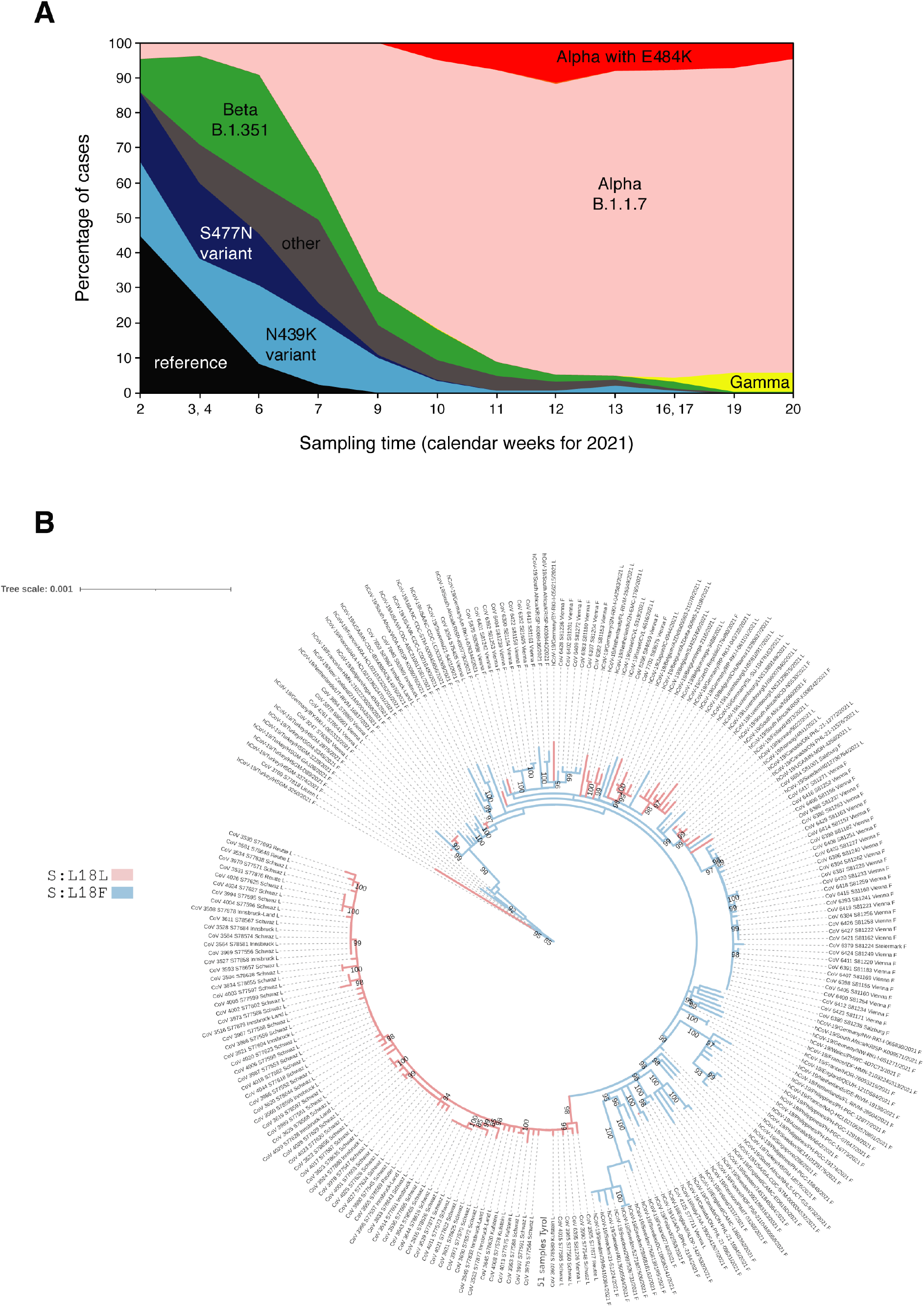
Timeline of observed SARS-CoV-2 strains in Tyrol. **A**. Timeline of identified strains based on samples analyzed in the period of January to May 2021. Results are plotted by calendar-weeks, weeks are joined if individual weeks consisted of <200 datapoints. For absolute numbers refer to Supplementary Table1, total number of samples analyzed is 8,426. Note the early coexistence of Alpha, Beta, other variants, as well as the reference strain. While Beta initially expanded in Tyrol, it was, like all other sub-strains, overtaken by the relative expansion of Alpha. **B**. Phylogenetic tree of Beta (B.1.351) in Austria based on whole genome sequencing. International cases are added to put distances in perspective. The position L18 separates the relatively homogenous cluster in Tyrol (L18L, blue) from the majority of cases outside Tyrol (L18F, red). Based on the data in this analysis no spread of the Beta cases of Tyrol to other countries is evident.

The unique and large outbreak of Beta in the state of Tyrol, that is geographically somewhat separated from the rest of the country (**Supplementary Fig. 6A**), allowed us to estimate national versus international spread of this variant across Austria. Interestingly, the Beta from Tyrol was distinct from that observed in Vienna and other parts of the country: non-Tyrol clusters typically contained the additional L18F mutation that was practically never observed within Tyrol (**Supplementary Fig. 6B**). We leveraged the higher coverage of the SARS-CoV-2 genome by WGS for in-depth lineage analysis of the Beta cases across Austria, generating a total of 162 sequences of Beta samples from Tyrol and 122 from the rest of the country. Our phylogenetic analysis showed that the Beta cluster in Tyrol formed a monophyletic clade. Unlike Beta variant sequences from other Austrian provinces than Tyrol, these samples did not harbor the Spike:L18F mutation. In contrast, Beta variant sequences from Vienna formed a number of separate clades dispersed across the global phylogenetic tree of this VOC, that are well separated from the Tyrol clade. Similar to the observations in Beta, we observed the mutation S640P appearing in the R.1 background within Vienna and in fact replacing the original R.1 strain, but we observed no spread of this sub-strain to Salzburg (**Supplementary Fig. 6C**). Distinct cluster fingerprints in different parts of the country highlight that even under the imposed travel restrictions international import of new VOCs into Austria remained frequent. This is corroborated by several distinct imports of Gamma and B.1.617 (Delta and Kappa) to Austria, a country of just 8.9 million people.

The pattern of increase in incidence of Alpha observed in Tyrol, mirrors the scenario observed across the world, and is a result of the markedly increased infectivity of this variant strain. The Alpha variant contains ten amino acid changes in critical domains of the spike protein relative to the reference, and thus provides a new starting point on which new mutations can accumulate. Given this modified starting point and the fast spread of Alpha, it is possible that mutations arising within an Alpha backbone will generate a further accelerated viral spread in the future. We therefore studied the mutational landscape within Alpha in more detail and found a staggering number of 160 distinct, fixed mutations or combinations of mutations in the S gene within the locally-limited patient cohort we analyzed (**Fig. 6A**). Given the complex structural changes the protein undergoes and the strong selective pressure, this degree of flexibility was surprising to us. Only few mutations were seen at a relatively high frequency and remained present over several weeks, while most others appeared sporadically. Numbers of more frequent mutations also fluctuated strongly likely due to local spreading, but we have not yet observed a sustained expansion for any of the mutations in the Alpha backbone. Interestingly, the most frequent mutation was found in position 716, where Alpha already displays a T>I exchange relative to the reference. We found that in 6.6% of Alpha in Austria, this was further mutated to valine. This is one of two incidences we observed of a second mutation occurring on an already variant codon, i.e. ACA(T)>ATA(I)>GTA(V). The other incidence is also in position 716 and produces a change to leucine, also most likely arising from an intermediate isoleucine codon. While these particular I>V or I>L replacements likely represent a minor change in biochemistry, they highlight the fact that such mutated backgrounds open new possibilities for the virus to explore in terms of available amino acid changes that can be reached with a further single-nucleotide substitution. Specifically, whereas the reference strain could change T716 to A, I, K, P, R and S with a single substitution, the Alpha strain can change I716 to K, L, M, R, T and V.

**Figure 6.**
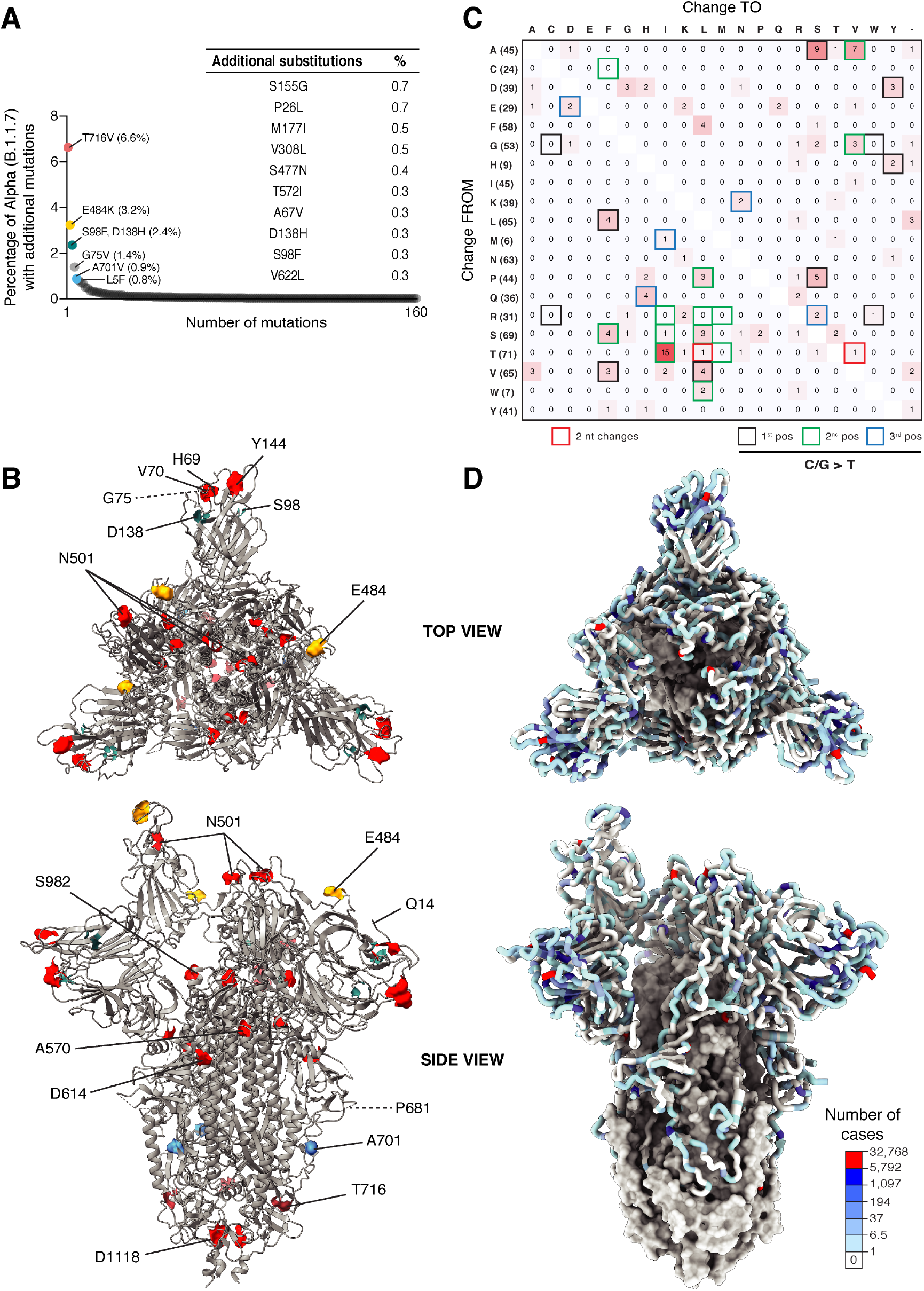
Mutational landscape of the spike protein. **A**/ Most frequently observed mutations or combinations of mutations in the Alpha (B.1.1.7) background. T716V represents a second-degree mutation away from T716I already mutated in Alpha. While few mutations were found frequently, most appeared in few patients and time- and space-restricted. **B.** Positions with mutations defining the Alpha backbone are labeled in red on structure of the Spike trimer with one open RBD (PDB: 6zgg). Frequently observed additional mutations are highlighted in colors matching panel A and labeled. Among them, E484K is located in a flexible loop that interacts with ACE2. **C.** Amino acid changes observed within the spike protein. Numbers refer to the count of positions with respective change, total number of each amino acid within the domain is shown in brackets next to the reference amino acid. Due to the restrictions imposed by the triplet code only few amino acid substitutions are observed. In two cases, a combination of two mutations within a codon lead to the observed change (red frame). The majority of changes is explained by a mutation of cytosine or guanine to thymidine, the respective codon position that is mutated is indicated with black, green, and blue frame respectively. **D** Mapping of all mutations observed in the Austrian surveillance dataset (Jan-Jun 2021) onto the spike crystal structure in open position with one RBD rotated upwards (PDB: 6zgg). Regions not covered by sequencing are shown as surface model in gray, while covered regions are in cartoon form. Color code represents how many cases of mutations were observed, independent of coexisting mutations. Positions mutated in the Alpha variant, and D614G are highlighted in red. Highest mutational frequency was observed on the protein surface, in particular on the distal surface of the NTD and within the ACE2 binding interface.

To gain further insight into where in the spike protein additional mutations occurred, we visualized the six most frequent ones on the protein structure^38^ (**Fig. 6B**). The second most frequent mutation in the B.1.1.7 backbone was E484K, which is located in the RBD and seems to change an epitope important for antibody-mediated neutralization. Intriguingly, the next most frequent mutation is a double substitution, S98F and D138H, both of which occur in the lobe formed by the N-terminal domain. This lobe already contains three amino acid deletions in B.1.1.7 (H69del, V70del, Y144del), and is also the region where the two of the next most frequent mutations occur, G75V and L5F. It is clear that this domain of the protein remains particularly flexible to change, and given that it represents an epitope for binding of neutralizing antibodies^39^, it should be monitored closely.

### Meta-analysis reveals mutational constraints and flexibility across the spike protein

To more generally explore the mutational landscape of the spike protein, we performed a meta-analysis on ~30,000 high quality sequences collected across Austria. We summarized all amino acid exchanges within the spike protein in our sample cohort, independently of their frequency across codon positions. Expectedly only a few substitutions were observed, reflecting primarily single-nucleotide changes (**Fig. 6C**). This is most likely due to the relatively low mutational rate of SARS-CoV-2 that limits the codon space the virus can explore in a given generation. A few amino acid changes were observed at particularly high frequency, with T>I being the most frequent. We thus also counted how often each type of nucleotide substitution is observed (**Supplementary Fig. 6D**). Exchange of C or G for U was observed at much higher frequency than others. This phenomenon had been previously described^40–43^ and is likely a cause of the U-biased genomes in Coronaviridae^44^. Interestingly, all frequent amino acid substitutions, and 58% of all substitutions (80/139), are explained by mutation of C or G to U, i.e. just two of the 12 possible exchanges. We propose that this mutational bias imposes a strong evolutionary constraint on SARS-CoV-2, limiting the number of observed amino acid changes and thus extending the time frame for the virus to explore a more infective state.

Several mutations frequently observed in the Alpha backbone were also observed in different backgrounds and in various contexts. Examples of such mutations are L5F, L18F, S477N, E484K, N501Y, P681H. This argues that at least for some positions, signs of converging evolution can be observed, an important consideration for example for development of second-generation SARS-CoV-2 vaccines. Of the 795 amino acids analyzed, we found mutations in 178 positions in the Austrian dataset (**Supplementary Table 4**). Of note, all 22 known glycosylation sites were excluded from mutagenesis. We also never observed mutations from or to cysteine in this sample set. A notable exception of this was seen recently in the Gamma lineage in two samples, where we found two mutations to cysteine, namely G199C and G232C. Intriguingly, these amino acids reside directly next to each other a n d t h u s c o u l d f o r m a d i s u l f i d e b r i d g e (**Supplementary Fig. 6E)**. We expect this might impact the structural stability of the NTD at the interface with the RBD.

The 178 mutated positions we identified are not randomly distributed but rather form clusters. Supporting this point, if a given amino acid is mutated, the likelihood of a change in one of its direct neighbors is 3.1 times higher (0.84 changes versus 0.27). We thus mapped the mutational flexibility of the spike ectodomain on the three-dimensional structure of the protein (**Fig. 6D**). It immediately became apparent that mutations cluster in the distal part of the NTD (mutations or deletions occur in practically all positions, e.g. 138-144 and 152-157) while the more proximal beta-sheet is mutated at much lower frequency. A high frequency of mutations is also found in the flexible part of the RBD spanning positions 475-478 and 483-484. The high throughput sequencing enabled by SARSeq S-tiling thus confirms observations on the evolutionary freedom of the SARS-CoV-2 spike protein from global datasets but also highlights that at least on a national level observed changes remain restricted and leave a large evolutionary space unexplored.

## Discussion

A functional and cost-effective tool for global surveillance of SARS-CoV-2 variants needs to fulfil several key parameters. It must be robust across all amplicons and samples, as well as sensitive enough to deliver across viral titers spanning a range of 10^6^ (Ct 13-33). Scalability is also important during a pandemic, as an effective surveillance strategy may require analysis of thousands of samples. This requires methods that can achieve high throughput while minimizing processing time and effort, reagent use and overall cost. Most NGS library preparation protocols, in particular those for targeted sequencing approaches, typically require one or more DNA purifications steps. While DNA purification can be parallelized and automated by use of magnetic beads, it still requires multiple pipetting steps, reagents and plasticware and thus extended processing time and effort. Here, we adapted SARSeq, a method that has proven robust and sensitive across multiple amplicons to achieve high-throughput and cost-effective sequencing of the SARS-CoV-2 S gene. SARSeq represents an “addition only” assay, it consists of simple pipetting steps where reagents or samples are added, and it does not require DNA purification steps. Moreover, all sequencing adapters are introduced during PCR so it bypasses the need for library preparation. SARSeq S-tiling maintains exquisite sensitivity, with 93-99% of amplicons detected from samples with a Ct range of 14-35.

Due to the focused sequencing of the spike gene ectodomain and the relatively even coverage of all amplicons, the sequencing depth required for SARSeq S-tiling is low and >20,000 samples could be multiplexed into a single sequencing run, bringing sequencing costs to a few cents per sample. A downside of the focused sequencing in the current version of SARSeq S-tiling, is the fact that it does not cover the entire sequence used in immunization approaches and therefore important mutations conferring immune evasion may be missed. While no such mutations are described to date, an extension of the covered region by addition of amplicons could be a useful add-on to the method. The limited coverage has also proven insufficient for robust lineage identification using the Pangolin package^36^. Lineage calling from S-gene tiling was therefore done by clustering of identical mutational profiles followed by human annotation. While this method is the best way to detect interesting mutational patterns early on, it is also time consuming and can certainly be further streamlined.

The simplicity and robustness of SARSeq S-tiling enabled us to set up a spike-gene, mutational surveillance pipeline for Austria in just two weeks. With the minimal requirement in pipetting steps, no complicated robotic setup is needed. With a team of 4 people, a dozen PCR machines and one liquid handling device we could ensure a throughput of 2304 samples/week including sample logistics and reporting. Even without any robotics, processing of ~ five hundred samples per person/per day is feasible using a multichannel pipet. The price per sample was ~15 EUR including sequencing but excluding RNA preparation. SARSeq S-tiling thus represents a scalable and easy to implement tool that allows for surveillance of pathogens at high throughput and minimal costs. In Austria, the pipeline we set up ensured surveillance of 10-50% of all SARS-CoV-2 infections in the first half of 2021 and could thereby inform health authorities to take adequate measures.

SARSeq S-tiling first detected outbreaks of multiple VOCs in the state of Tyrol, namely Alpha, Beta, Gamma and Delta. In particular for Beta measures were taken to contain its spread through quarantine of Tyrol by border control and obligatory testing for exit. In addition a prioritized vaccination program in the most affected region was set up. The Alpha (B.1.1.7) variant overtook not only the original strains previously present in Tyrol, but also all other VOCs. Its earlier prevalence in the rest of Austria resulted in stronger local containment measures, which likely contributed to prevent the quantitative spread of Beta into these regions. This confirms that the best way to prevent the spread of a variant is by increased containment measures such as masks^45^, albeit ideally imposed before a new and more contagious variant is expanding. The observation in Tyrol is to our knowledge a unique situation, in which the relative fitness of these four VOCs could be assessed within a relatively confined geographic location and under the same measures for viral restriction.

While Beta, Gamma, Delta originated from B.1 upon fixation of the D614G mutation on the reference strain, this original backbone is practically extinct in most parts of the world. Therefore all further VOCs with further complex changes will originate from the backbones that are currently prevalent, in particular Alpha and Delta. Such second-generation lineages have the potential to explore amino-acid changes not available to the reference strain and therefore represent an unknown risk. Early detection of such new lineages requires intensive sequencing efforts and close inspection of the changes with a focus on the S gene. We thus performed meta-analyses of the mutational landscape of the spike protein. This revealed a number of positions that change at high frequency, whereas most positions showed no change in our dataset of ~30,000 high-quality sequences. The majority of amino acid changes were caused by C or G mutations to U. This mutational bias at the nucleotide level skews the frequency and position of observed amino acid changes. Many such amino acid substitutions may be under purifying selection and thus not manifested in our dataset (**Fig. 6B**), a few others seem to be either neutral or beneficial and are frequently observed in multiple different backgrounds (i.e. are highly homoplasic). Since amino acid changes happen typically by single nucleotide exchanges, every amino acid can only convert to few others. For example, E484 is converted to K by an exchange of G>A in the first position of the codon in the Beta strain, and to Q by G>C mutation in Kappa (B.1.617.1). From both, an A>G exchange in the second position of the codon could now result in R484, which has experimentally been shown to most strongly enhance receptor binding^46^. Most amino acids changes likely still remain unexplored, but can be reached by a second mutagenic step. If the initial mutation is not favored or even disfavored however, that second step remains rare. Indeed, we only observe 2 cases of such two-step transitions among the total of 237 distinct amino acid substitutions occurring in 178 positions within the spike protein. Together with the strong bias towards mutations to U the evolutionary freedom of SARS-CoV-2 thus remains somewhat restricted, but will be explored eventually if the global viral population continues to be high. Depending on the incidences of SARS-CoV2 infections in the years ahead, dense surveillance might continue to be a necessity. S-gene tiling by SARSeq represents an ideal platform to enable this in a cost-effective and scalable way.

## Materials and Methods

### Sample material and ethics

This study involves the use of patient medical data from EMS (Epidemiologisches Meldesystem), which is the official source for public health surveillance activities in Austria. EMS follows the mandate of Austrian and international legislation and is hosted by the Austrian Agency for Health and Food Safety (AGES). The present study was approved by the local Ethics Committee of Vienna (#EK 21-141-0721) and the Federal Office for Safety and Health Care (BASG). Sequencing and analyses of virus whole genomes were approved by the Ethics Committee of the Medical University of Vienna, Austria (2283/2019). The study was conducted in accordance with the Declaration of Helsinki.

### RNA purification

Left-over, purified RNA samples from SARS-CoV-2 positive samples was provided by the Austrian Agency for Health and Food Safety (AGES) and remained fully anonymized to us. These had been purified by various laboratories using different purification pipelines and were shipped regularly.

### Reverse transcription

For each sample, RT and and PCR1 are performed in two independent reactions, one for Odd amplicons and second for Even amplicons. Per sample 5 µl of RNA sample was mixed with 20 µl of reverse transcription reaction master mix. RT reaction master mix is prepared by combining 13.8 µl of nuclease free water, 2.5 µl of 10X RT buffer, 0.5 µl of dNTPs (25 mM each), 0.1 µl of 1M DTT, 2 µl of RT primer mix (Odd set contains 8 primers and Even set contains 6 primers. Sequences of RT primers can be found in Supplementary Dataset 1), 0.5 µl of Ribonuclease inhibitor and 0.5 µl Reverse transcriptase. RT mix was arrayed out in 96 well plates, stored on ice, and reactions were started within 1h. The RNA sample and RT reaction master mix combination was mixed 10 times by the liquid handling robot and incubated in a humid incubator at 55°C for 40 min. Subsequently the reverse transcriptase was inactivated at 95°C for 3 min and the reaction cooled down to 4°C. Supplementary Data 1 provide detailed concentrations and calculations on volumes of the pipetting scheme. For regular analysis components were premixed and frozen in aliquots such that upon addition of 15ml ddH_2_O and enzymes the mix was ready for 12 x 96-well plates (see Supplementary Data 3).

> 10× RT Buffer:
>
> 200 mM Tris-HCl pH 8.3
>
> 500 mM KCl
>
> 50 mM MgCl_2_
>
> 200 mM (NH_4_)_2_SO_4_
>
> 1% Triton X-100

### First PCR (sample indexing)

The PCR top-up master mix was prepared by combining 11.43 µl of nuclease free water, 2.5 µl of 10X PCR top-up buffer, 0.07 µl of dUTP (100 mM stock), 0.5 µl of Antarctic thermolabile UDG, 0.5 µl Hotstart Taq Polymerase and 10 µl of PCR primer mix with sample specific unique dual indexes (ODD set contains 7 primer pairs/1 µM each OR EVEN set contains 6 primer primers/1 µM each, sequences of PCR primers can be found in Supplementary Data 1). 25 µl of PCR top-up master mix per reaction is added and mixed ten times with the liquid handling robot. The reaction should be kept on ice. The mixed reaction is removed from ice, kept at RT for 5 min to allow reaction of the UDG, and put into a thermocycler that is already at 95°C (hot start). The program is: 95°C for 3 min, 10 high stringency PCR cycles (95°C for 30 sec and 63°C for 5 min) followed by 35 high efficiency cycles (95°C for 30 sec, 63°C for 30 sec and 72°C for 40 sec), then cool down to 12°C. Successful amplification of S-gene specific amplicons (~330bp) can be analyzed by resolving 10 µl of PCR reaction on a 2% agarose gel.

A master mix containing all components listed below, including homemade HotStart Taq Polymerase and Uracil DNA glycosylase (Antarctic Thermolabile UDG from NEB) was prepared and distributed to a 96-well plate containing previously arrayed primer pairs (multiple primer plates can be prepared simultaneously and stored frozen at −20°C). Using a liquid-handling robot, the primers and PCR master mix were mixed thoroughly and 25 µL of this complete 2× PCR mix were added to the 25 µL RT reactions ran as described above. Plates were sealed with aluminum sealing foil and incubated in a thermocycler following the conditions listed below.

All components were kept at room temperature during reaction set up and for 5 min before starting the PCR program, this provides the right conditions for UDG to act on Uracil-containing amplification products of previous PCR reactions, thereby removing spurious carry over contaminants. After UDG heat inactivation, the subsequent PCR reaction was again carried out in the presence of UTP to prevent carry over contamination in following runs. Supplementary Data 1 provide detailed concentrations and calculations on volumes of the pipetting scheme. For regular analysis components were premixed and frozen in aliquots such that upon addition of 15 ml ddH_2_O and enzymes the mix was ready for 12 x 96well plates (see Supplementary Data 3).

> 10× PCR Top Up Buffer:
>
> 750 mM Tris-HCl pH 8.3
>
> 200 mM (NH_4_)_2_SO_4_
>
> 1% Triton X-100

### Plate pooling

All well-barcoded PCR products from each row of a 96-well plate were pooled, typically 10 µL of each reaction were combined in an 8-tube PCR strip using a multi-channel pipette, and after mixing thoroughly, 50 µl of each the ODD and EVEN pooled mixes were transferred to a new 96-well plate (see schematic in **Supplementary Dataset 2**). This was repeated for every pair of PCR plates. 5 µL from each plate pool were re-arrayed in a new 96-well plate and treated with 2 µL of illustra ExoProStar 1-step for 30min at 37°C followed by 15 min at 80°C to remove any left-over primer.

### Second PCR (plate indexing and addition of sequencing adaptors)

A master mix with all components listed below was distributed across a 96-well plate (40 µL/well). To each we added 7.5 µL of unique dual-indexed i5/i7 primer pairs (Custom synthesized index primers with Nextflex barcodes, arrayed in 96-well plates) and 2.5 µL of ExoProStar-treated PCR1 pool. The reactions were run for 8 cycles to add sequencing adaptors with plate barcodes. 2.5 µL of ExoProStar-treated PCR1 pool are transferred into a 96-well plate containing and array of unique dual-indexed i5/i7 primer pairs (7,5ul each, custom synthesized index primers with Nextflex barcodes). To avoid cross contamination from previous runs indices used for PCR 2 were alternated between weeks. A master mix with all components listed below was added to each well. The reactions were run for 8 cycles to add sequencing adaptors with plate barcodes.

10x Sequencing-ready PCR Buffer:

> 750 mM Tris-HCl pH 8.3
>
> 200 mM (NH_4_)_2_SO_4_
>
> 20 mM MgCl_2_
>
> 0.1% Tween 20
>
> Master mix composition per reaction/well (volumes in µL):

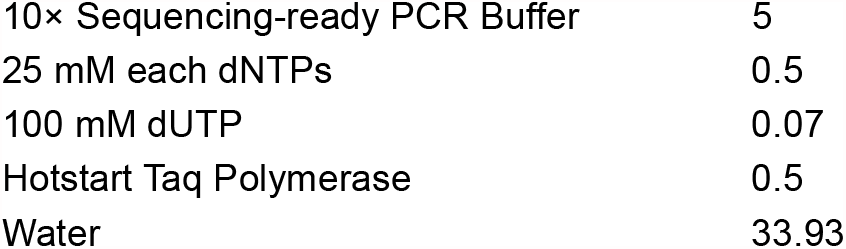

> Thermocycler program:
>
> 3 min at 95°C
>
> 8 cycles of: 30 sec at 95°C, 30 sec at 65°C, 40 sec at 72°C
>
> 3 min at 72°C
>
> Cool down to 12°C

### Final Pooling and preparation for sequencing

All samples from a 96-well plate (20µl from each well) were pooled and 200µl of pooled sample was resolved on a 1,5% agarose gel and 400-500 bp amplicons were excised and gel purified using Qiagen gel extraction kit.

### Sequencing

Quality control of libraries was performed in the fast, optimized manner described previously^29^. Libraries were sequenced on a NovaSeq6000 SP flowcell with a loading concentration of 220 pM and a custom read mode of 156/9/9/164 (read 1/index1/index2/read2). To increase the complexity of the sequencing sample, 10 % PhiX were spiked-in in every run. Due to the combinatorial two-dimension barcoding of the libraries, bleach washing steps on the NovaSeq can be omitted, because DNA remnants, potentially present in the sequencer’s fluidics from previous runs, can be filtered out. Nevertheless, a bleach wash was performed every 14 days.

### Data analysis

Paired-end (PE) reads were assigned to sample IDs using the two-dimensional dual indexing as described (SARSseq^29^). Subsequently, the PE reads of each sample were further assigned to amplicons/tiles based on the respective amplicon/tile-specific primer sequences that are contained within the reads using a custom awk script, enabling the specific alignment of each amplicon/tile. The PE-read-to-tile alignment was performed using minimap v2.17 and the SARS-CoF-2 reference sequence MN908947.3 from which we extracted the corresponding tile/amplicon reference sequences. After alignment, we retained only reads that mapped in proper pairs using samtools v1.10. a textual pileup of the filtered alignment was obtained using samtools mpileup 1.10 using parameters “-aa -- max-depth 0 --no-BAQ --min-MQ 1” and further summarized using readstomper.pl and R v3.6.3. We derived a sample-specific S gene consensus sequence from the pileup and translated this consensus sequence into the corresponding S protein sequence. The consensus encoding was based on two cutoffs-a minimal read cutoff (mRC) of 10 and a per tile confident coverage cutoff (cRC) defined as 1% of the median observed coverage of that tile in the respective experiment. Using these cutoffs we created an annotated consensus sequence highlighting variants to the reference according to the following rules: ““: read coverage < mRC; “_”: read coverage < cRC and no variant allele above 0.5; “lower case” or “.”: read coverage < cRC and variant allele/deletion above 0.5 or confident read coverage and variant allele observed in 0.2-0.5 fraction of reads; “upper case” or “-”: variant allele or deletion with confident read coverage.

### S-gene specific amplicons

Suppl. Data 1, tab “Amplicons” contains all amplicons that were used for tiling of the SARSeq S-gene. Primer sequence is highlighted in green (forward primer) and red (reverse primer) in each amplicon and provided individually in 5’ to 3’ orientation. The region extracted for analysis is highlighted in bold in the amplicon and also provided separately.

### Whole genome sequencing

We followed COVID-19 ARTIC v3 Illumina library construction and sequencing protocol V.4, as previously described^15^. Shortly, cDNA was reverse-transcribed from isolated viral RNA using Superscript IV Reverse Transcriptase (ThermoFisher) and used as a template to amplify overlapping viral fragments using the ARTIC Network initiative version 3 primer set^47^. The pooled PCR product were cleaned with AMPure XP beads (Beckman Coulter) and amplicon concentrations and length distributions checked with the Qubit Flourometric Quantitation system (Life Technologies) and the 2100 Bioanalyzer system (Agilent). Libraries were created using the NEBNext Ultra II DNA Library Prep Kit for Illumina (New England Biolabs) and sequenced on the NovaSeq 6000 platform (Illumina) in 250 bp paired end mode using an SP flowcell.

After demultiplexing, adapter sequences were trimmed from reads using BBDUK and overlapping sequences of each readpair were corrected using BBMERGE, both from the BBtools suite (https://jgi.doe.gov/data-and-tools/bbtools)^48^. BWA-MEM (v. 0.7.17)^49^ was used to map read pairs were to both the human (Hg38) and the SARS-CoV-2 genome (GenBank: MN908947.3, RefSeq: NC_045512.2) with a minimal seed length of 17. Only reads mapping uniquely as proper pairs to the viral genome were retained and primer sequences were removed by soft-clipping with iVar^50^. Mapped reads were then downsampled to at most 10000 read pairs starting at any given position and realigned using the Viterbi method provided by Lofreq (v 2.1.2)^51^. Samtools mpileup (v 1.9)^52^, bctfools (v 1.9)^52^ and SEQTK (https://github.com/lh3/seqtk) were used to create a consensus genome from the aligned reads using a minimal coverage of 15.

### Quantitative RT PCR (qPCR) assay

The RT was performed as described in the reverse transcription section, using 5 µl of sample in 25 µl RT reaction. For qPCR analysis, 25 µl of PCR1 top up reaction mix with 1.5 µl of CDC-N1 primer/probe set (IDT 10006713/sub part 10006600) was added to the completed 25 µl RT reaction mix. Reactions were run at 95°C for 3 minutes, followed by 45 cycles of 95°C for 15 seconds and 55°C for 45 seconds in a BioRad CFX Connect™ Real-Time System.

## Generation of figures

Statistical analysis and plots were done using GraphPad Prism 8.4.3., plate layouts were illustrated in Microsoft Excel, and figures were assembled in Adobe Illustrator CS6.

## Code availability statement

Custom code was used to analyze all NGS data. The script is available at GitHub under https://github.com/sarseq/sarseq2.

## Acknowledgements

We thank all the diagnostic labs in Austria that provided SARS-CoV-2 positive patient samples. Our gratitude goes to “Matt” James Watson for support with all contractural issues and IMBA and IMP management for strongest support of the pipeline, specifically Jürgen Knoblich, Harald Isemann, Markus Kiess, Jan-Michael Peters and everyone else involved. We are grateful to Stefan Ameres, Julius Brennecke, Harald Isemann, Andrea Pauli, Johannes Zuber for close interaction throughout. We are indebted to David Drechsel and his team as well as Robert Heinen, Kristina Uzunova, Tim Clausen and Anton Meinhart for supporting enzyme purification efforts, to Andreas Sommer and the entire NGS team at the VBCF. We thank Clemens Plaschka for help with structure visualization and generation of publication-quality figure. Last but not least we would also like to thank our labs and all coworkers for understanding and supporting us in so many ways. R.Y. was partially supported by Oliver Bell’s laboratory at USC. Curiosity driven biomedical research at the IMP is largely sponsored by Boehringer Ingelheim. IMBA is generously funded by the OEAW.

The current pandemic has challenged healthcare systems worldwide, but it has also led to an unprecedented concerted response on multiple levels. We wish to contribute to the fight and therefore welcome anyone interested in implementing this method to contact us.

## Author contributions

E.Ö. and M.M.S. prepared and managed reagents for the high-throughput pipeline, processed samples with help from R.Y., T.A., T.P., and V.F., and participated in data analysis. M.N. set up the bioinformatic analysis pipeline with guidance by A.S. and performed data analysis for figures. R.Y. supported development of the method and the analysis pipeline. P.T. and L.E. performed phylogenetic analysis, T.P. processed samples for WGS. A.V. optimized the NGS workflow, I.T. set up the bioinformatic handover protocol. T.S. and M.F. provided advice and support for establishment of the clinical study and accompanying ethics. R.H. processed and supplied samples for the Tyrol analysis. A.I. supplied samples from various labs and supported annotation, D.S. supported epidemiological analysis. C.B. supervised WGS analyses, A.B. oversaw WGS and supported various aspects of the study. A.S. helped design the bioinformatic pipeline, provided guidance and support for its implementation, and helped write the manuscript. F.A. set up the surveillance program and ensured financial support. U.C. and L.E. developed the method, performed analysis, generated figures, and wrote the manuscript.

## Supplementary Figures

**Supplementary Figure 1.**
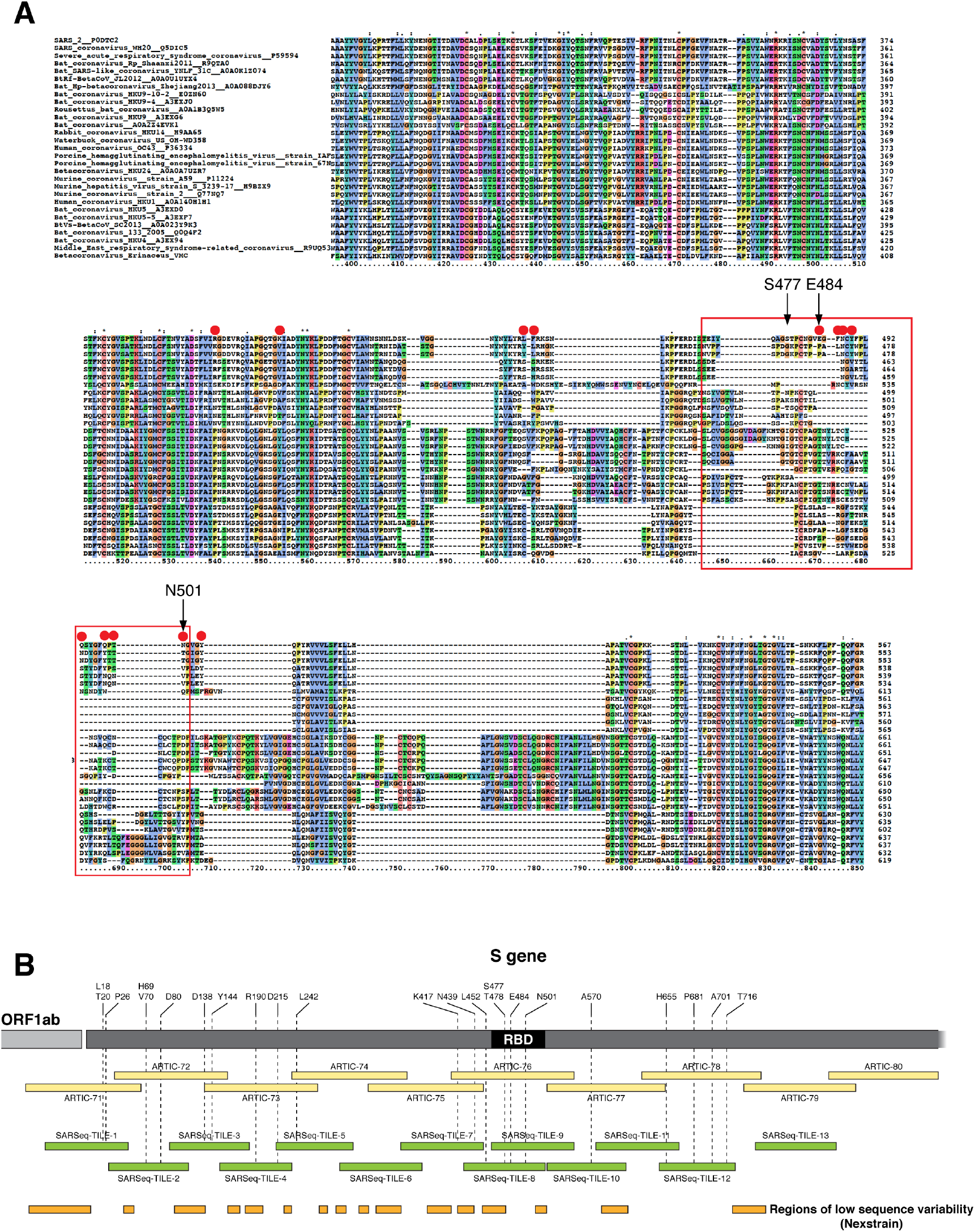
Design criteria for the SARSeq S-gene tiles. **A**. Multiple sequence alignment of SARS-CoV-2 (top row) and closely related coronaviruses. The highest degree of variability is seen in the region highlighted by a red frame. Insertions are also seen in some strains in the adjacent protein region. Amino acids interacting with the human ACE2 receptor are highlighted with red circles. Given the importance of this region for neutralizing antibody binding and infectivity it was decided to cover the variable part of the receptor binding domain highlighted with a red frame with two independent amplicons (tiles 8 & 9). **B**. Scheme showing the relative location of ARTIC and SARSeq amplicons in the spike coding sequence. ARTIC amplicons are typically 500 bp long, SARSeq tiles reach a maximum of 280 bp. The region from amino acid 470-502 harboring the important mutations S477N, T478K (Delta), E484K (Beta, Gamma), N501Y(Alpha, Beta, Gamma) is covered with two amplicons in SARSeq. Tiles were designed trying to place primer binding sites in regions of lowest sequence variability (based on Nextstrain data in December 2020).

**Supplementary Figure 2.**
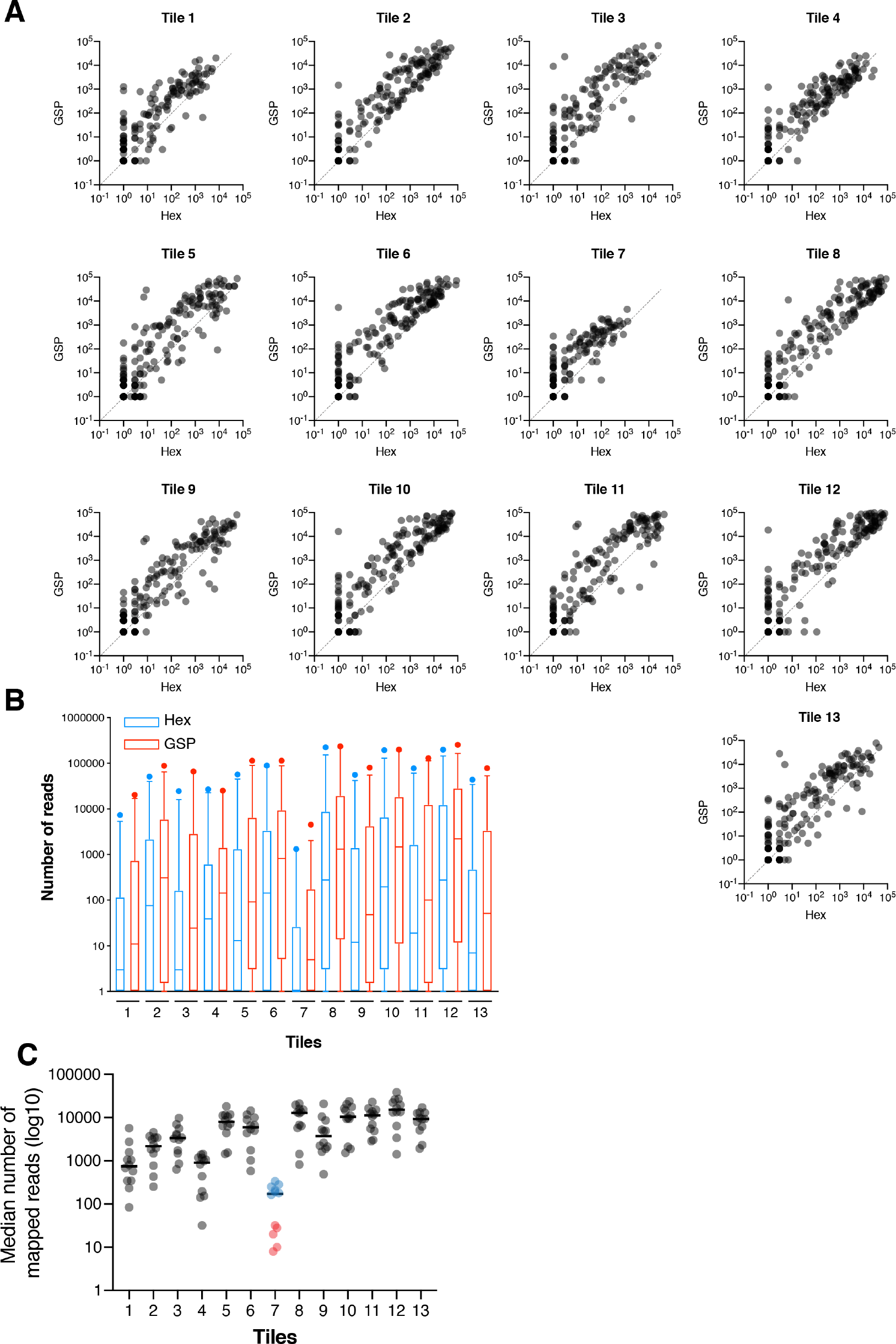
Effect of gene-specific priming during RT. **A**. Scatter plots of read depth obtained by SARSeq S-tiling upon reverse transcription primed either with random hexamers (X-axis) or S-gene specific primers (GSP) with a melting temperature of approximately 58°C. Each dot represents a clinical sample (n=192). For all tiles, gene specific primers resulted in higher coverage. Note that many samples were only covered with gene specific RT primers, this is particularly evident for amplicon 7 that typically reached lower read depth. **B**. Box plot summarizing data from A. **C**. Median number of reads obtained for each of the 13 tiles, per independent analysis run (each run consisted of ~2,300 samples). Tile 7 reproducibly generates the lowest number of reads, but read number increased about ten-fold when adding a second tile-specific RT primer (blue runs versus red runs).

**Supplementary Figure 3.**
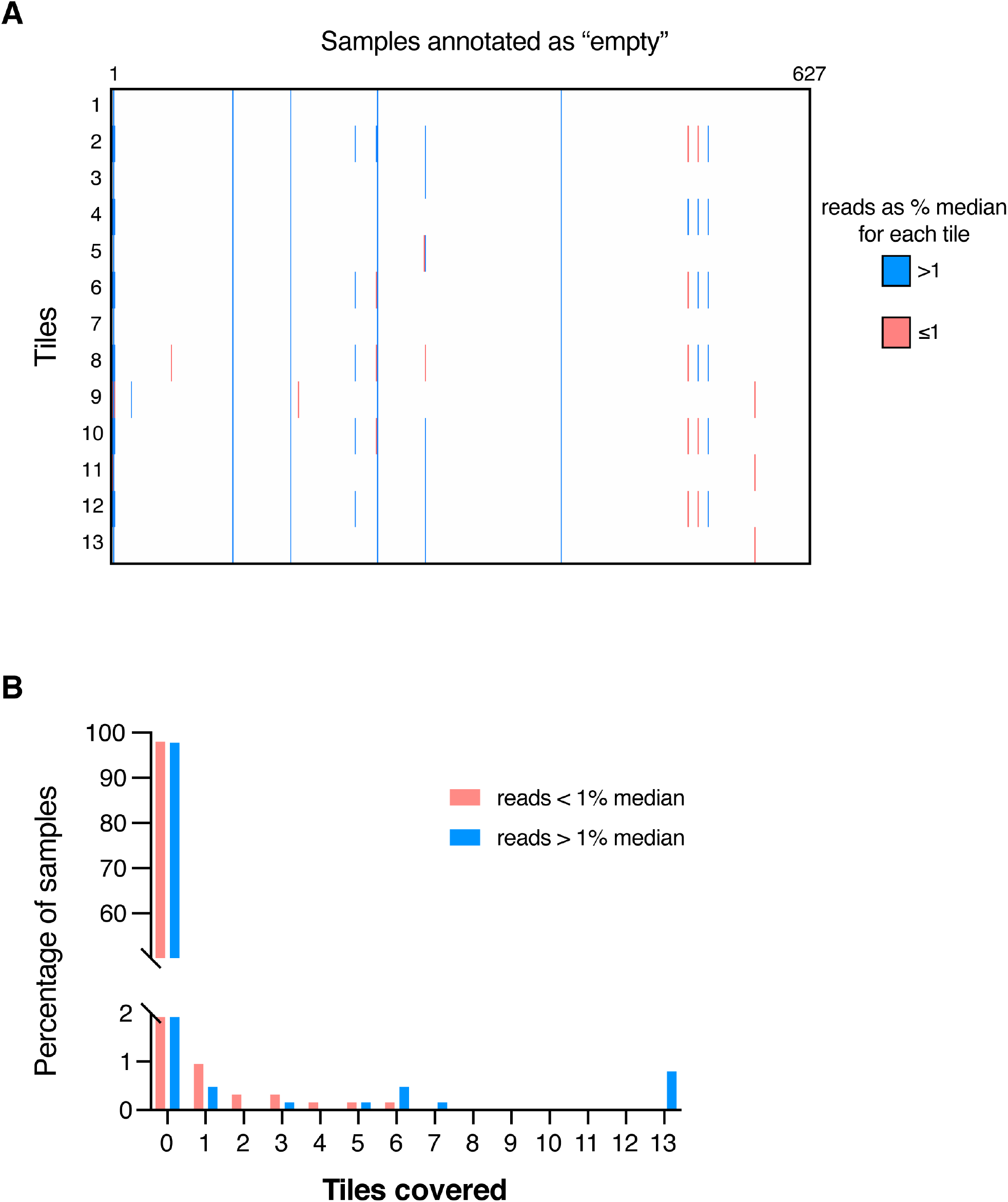
False positive rate. **A**. Heat map of reads per sample in 627 presumably empty wells (as annotated by clinical laboratories) from five independent processing/sequencing runs. The number of reads is represented in three categories: absence of reads (or reads <10) is shown in white; reads equal or greater than 10 but below the cutoff of 1% of tile-specific median read depth are shown in pink; tiles covered above the cutoff are shown in blue. Note that reads appear in a non-random pattern, i.e. unlikely to reflect contamination by one or few amplicons but rather suggesting that those positions are not truly empty of template RNA. **B**. Histogram summarizing coverage from dataset in A. While no signal can be detected for almost all samples expected to be empty, <1% of positions showed coverage of 1 or more tiles. The relatively high number of samples displaying reads in 6 or 13 tiles is likely due to experimental spillage or true positive positions and not explained by index hopping or another system-inherent error.

**Supplementary Figure 4.**
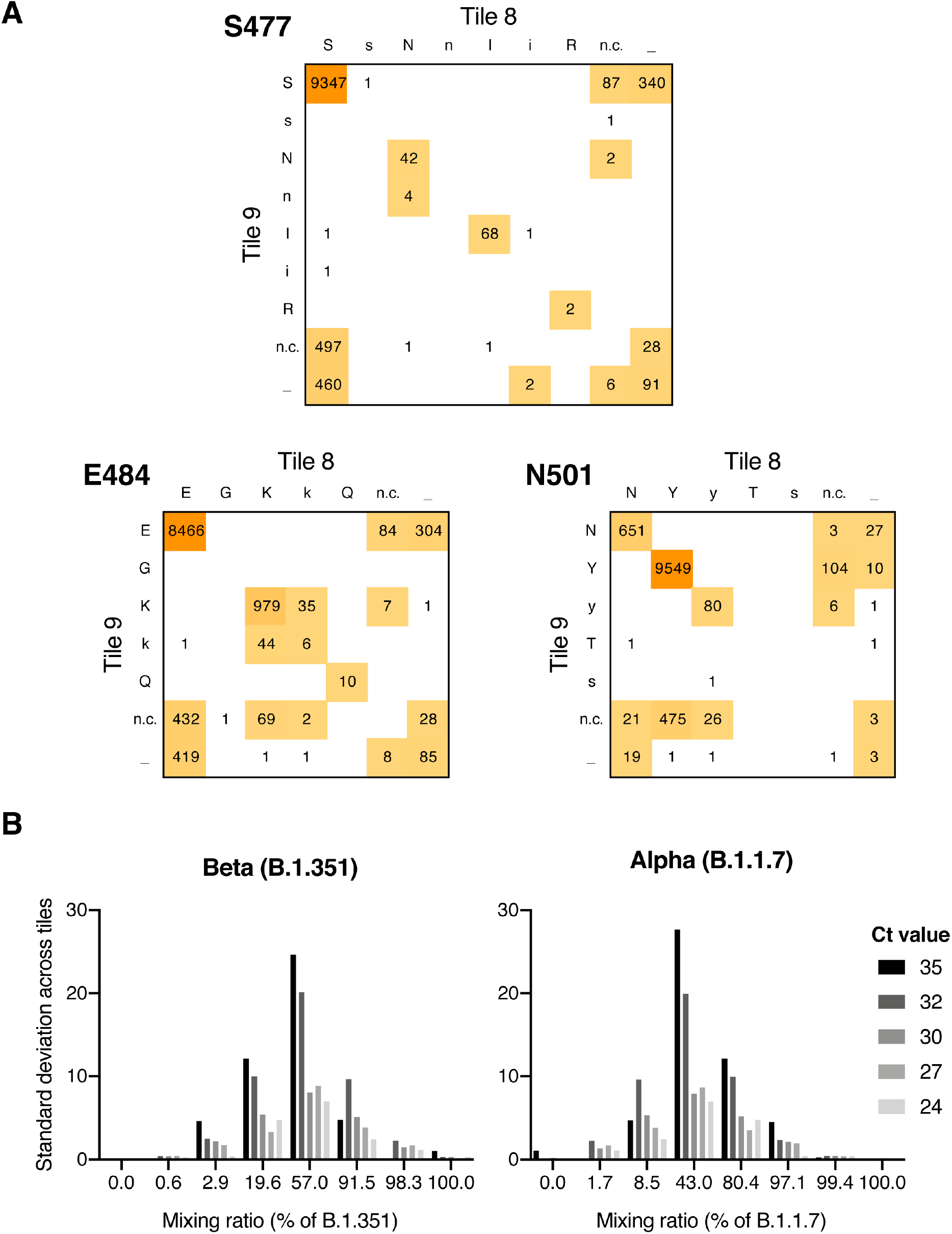
Reproducibility and quantitativeness of SARSeq S-tiling. **A**. Comparison of sequence consensus obtained from tile 8 and 9 across >10,000 samples, extension to Fig. 3A. While coverage is not always obtained from both tiles, existing results are highly concordant when both produce sequences. Note that amino acids denoted here as small letters can be due to low confidence (below cutoff of 1% median but above 10 reads, see main text) or low penetrance/fraction of reads within the sample (20-50% of reads contain that substitution). Therefore, the reproducible discrimination between upper and lowercase positions is not expected, it is merely an added information to the output of the bioinformatic pipeline. **B**. Standard deviation of data presented in Fig. 3D, E. As anticipated, error rates of quantitativeness per amplicon are higher for low titer/high Ct value across all ranges of mixing ratios. For Ct values <32 error rates reach a maximum of ~10% at equimolar mixing.

**Supplementary Figure 5.**
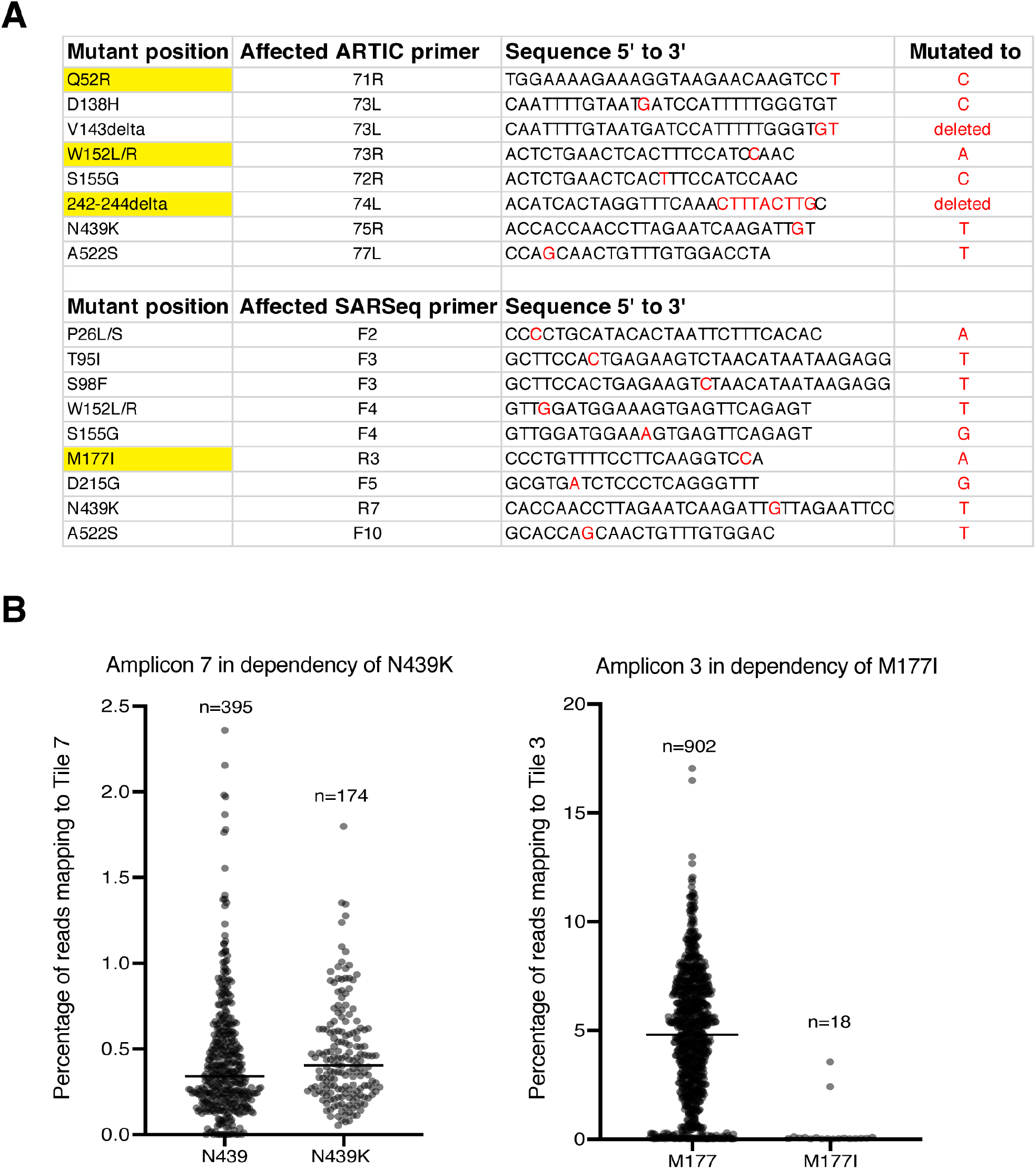
Effect of mutations on priming neighboring amplificons. **A**. Table showing all ARTIC primers and SARSeq primers that overlap with at least one of the 30 most frequently observed mutations in the Austrian SARS-CoV-2 surveillance cohort. Mutated nucleotides are highlighted in red. Yellow highlights show mutations with experimental evidence (Fig. 4 A, D and this figure, panel B) for perturbation of PCR, as expected based on mismatches close to the 3’ end. Mutations are anticipated to be particular inhibitory if close to the primer 3’ end. **B** Analysis of SARSeq amplicons 7 and 3 respectively in relation to presence of indicated mutations overlapping the primer binding sites. While the mutation resulting in the frequent N439K mutation is not affecting PCR, the rare M177I mutation is consistent with its 3’ proximity. Note that the N439K mutation also overlaps with the ARTIC primer 75R, albeit closer to the 3’ end.

**Supplementary Figure 6.**
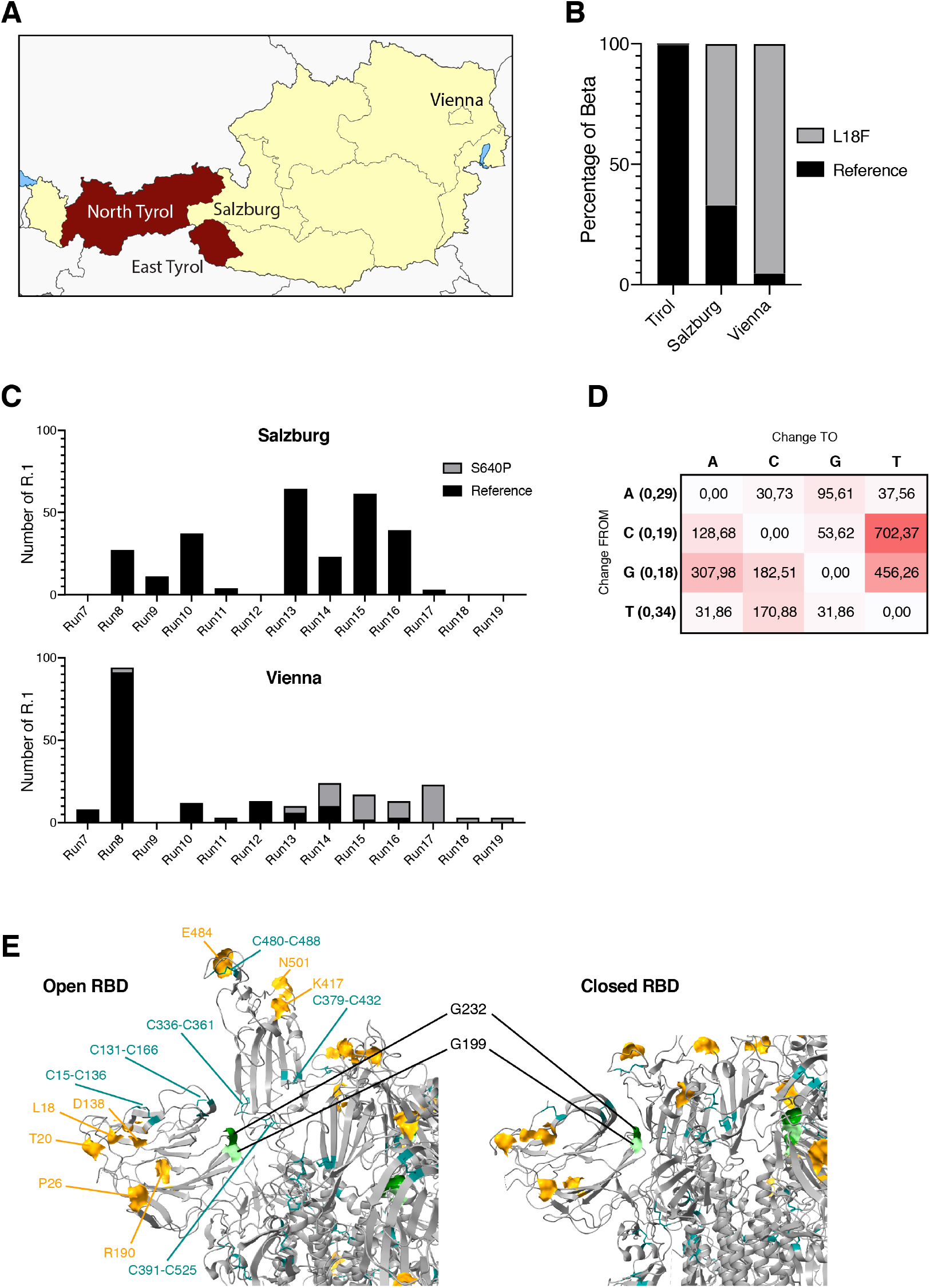
Mutational surveillance of Austria. **A**. Geographic overview of Austria. Tyrol is located in a western extension of the country and separated by mountains. Salzburg is a state neighboring Tyrol, while the capital Vienna is located in the east and thus further away from Tyrol. **B**. Frequency of the L18F mutation in the B. 1.351/Beta backbone in Tyrol, Salzburg, and Vienna. While cases of Beta infection in Tyrol exclusively displayed the reference mutation, L18F was the great majority in Vienna. Salzburg displayed a mixed image consistent with its geographic location and expected human mobility. **C**. Time course of identified incidence of R.1 in Salzburg and Vienna. Absolute numbers vary greatly between runs (one run per week) due to sampling depth of the different regions. While S640P appeared in Vienna in run 8 and eventually made up for all detected cases this mutation was never observed in Salzburg arguing for independent epidemiological clusters without exchange. **D**. Relative frequency of nucleotide changes within the covered region. While the coding sequence already shows a bias towards Thymidine (Uracil in the genome) (0.34 shown in brackets), nucleotide changes from cytosine or guanine to thymidine were the most frequent. **E**. Location of two Gly to Cys mutations in positions 199 and 232, which likely form a disulfide bond, in the backbone of Gamma (P.1). (Left) RBD in its open position. (Right) closed RBD conformation. The mutations are found at the proximal side of the NTD close to the interface with the RBD. Gamma-specific mutations are shown as yellow surface, all disulfide bridges are shown in stick format.

